# Polygenic prediction of fear learning is mediated by brain connectivity

**DOI:** 10.1101/2025.03.12.25323754

**Authors:** Javier E. Schneider Penate, Carlos A. Gomes, Tamas Spisak, Erhan Genc, Christian J. Merz, Oliver T. Wolf, Harald H. Quick, Sigrid Elsenbruch, Harald Engler, Christoph Fraenz, Dorothea Metzen, Thomas M. Ernst, Andreas Thieme, Giorgi Batsikadze, Bianca Hagedorn, Dagmar Timmann, Onur Güntürkün, Nikolai Axmacher, Robert Kumsta

## Abstract

**Background:** Genetic variants may impact connectivity in the fear network such that genetically driven alterations of network properties (partially) explain individual differences in learning. Our aim was to identify genetic indices that predict physiological measures of fear learning mediated by MRI-based connectivity.

**Methods:** We built prediction models using exploratory mediation analysis. Predictors were polygenic scores for several psychological disorders, neuroticism, cross-disorder risk, cognitive traits, and gene expression-based scores. Candidate mediators were structural and functional connectivity estimates between the hippocampus, amygdala, dorsal anterior cingulate, ventromedial prefrontal cortex and cerebellar nuclei. Learning measures based on skin conductance responses to conditioned fear stimuli (CS+), conditioned safety cues (CS-), and differential learning (CS+ vs. CS-), for both acquisition and extinction training served as outcomes.

**Results:** Reliable prediction of learning indices was achieved by means of conventional polygenic score construction but also by modelling cross-trait and trait-specific effects of genetic variants. A latent factor of disorder risk as well as major depressive disorder conditioned on other traits were related to the acquisition of conditioned fear. Polygenic scores for short-term memory showed an association with safety cue learning. During extinction, genetic indices for neuroticism and verbal learning were predictive of CS+ and differential learning, respectively. While mediation effects depended on connectivity modality, prediction of fear involved all regions of interest. Expression-based scores showed no associations.

**Conclusions:** Our findings highlight the utility of leveraging pleiotropy to improve complex trait prediction and brain connectivity as a promising endophenotype to understand the pathways between genetic variation and fear expression.

## Introduction

Classical fear conditioning is to date the most extensively employed paradigm to study fear learning experimentally. Typically, acquiring conditioned fear consists of repeatedly pairing a neutral stimulus to an aversive one, the unconditioned stimulus (US), when both are presented closely in time. After enough iterations, the neutral stimulus elicits a conditioned fear response by itself and becomes a conditioned stimulus (CS+). Extinction learning, on the other hand, is thought to result of a new association between the previously associated stimuli in which their contingency no longer holds and thus expression of fear to the CS+ reduces in magnitude and frequency (1). Moreover, it is commonplace to include stimuli that are never paired with the US in ‘differential fear learning’ protocols that serve as a control condition against which to measure the difference in conditioned responses to the CS+. Fear responses to these ‘CS-‘ alone are occasionally studied as measures of safety cue learning as well (2,3). Throughout this article we focus on the skin conductance response (SCR), since it is by far the most common operationalization of fear expression when studying human participants. Clinical populations show marked differences in their expression of fear and rate of fear and extinction learning compared to groups of healthy controls (4). Hence, alterations in fear learning mechanisms are thought to be involved in the aetiology of several mood-, anxiety, and stress related mental disorders. Although many studies support this notion, the experimental results it is based on are highly heterogeneous (2,3) and partly contradictory (5). This diffuse picture could be attributed to sample assemblage or the experimental design, which makes result comparisons across studies difficult, but also to an inherent variability between individuals that cannot be explained by neither noise nor statistical confounds. While the former is a widely acknowledged concern, the latter is still poorly explored (6). The present work is an effort to advance knowledge with respect to interindividual differences in fear learning and extinction.

Thanks to genome-wide association studies (GWAS) it has now become clear that a fraction of phenotypical variability in virtually all traits, including mental health, has its roots in genetic effects spread across a large number of loci over the entire genome (7,8). To make sense of this polygenicity, polygenic scores (PGS) have been proposed as a measure to harness the additive genetic effect on phenotypes. The approach involves aggregating the myriad of minute effects of single nucleotide polymorphisms (SNPs) into one single score that reflects the genetic liability towards a specific phenotype (9). Importantly, PGS are computed at the individual level, making the study of inter-individual differences possible. The genetic predisposition for various mental disorders and learning-related traits may have a nonnegligible impact on the expression of fear, even in the absence of symptoms of clinical relevance. Here, we put that idea to the test by assessing the predictive power of PGS in a pooled sample of healthy individuals.

Although their predictive capabilities have been shown, conventional PGS still lack an insight into the biological mechanisms that lie inevitably between the polymorphisms and the behavioral traits upon which they are constructed. More recently, the expression-based polygenic score (ePGS) has been proposed as an adaptation to bridge the two in a more mechanistic matter (10). They are built under the assumption that polymorphisms likely affect gene expression of one or several genes in near proximity; genes which are functionally coregulated by networks that work toward specific biological pathways (10,11). Here, we used transcriptomic summary statistics from the Genome Tissue Expression Project (12) (GTEx) to implement two network-building strategies. Firstly, we extracted co-expression modules containing the *DCC* (deleted in colorectal cancer) gene due to its expression being instrumental for neurotypical development, axonal guidance and growth, and the regulation of white matter tract organization (13,14). Secondly, we attempted to identify networks involved in Hebbian learning and plasticity. Gene modules that reflect those processes may leave a durable mark on the brain’s structure and function, which could in turn affect associative learning.

Recent work uncovered an extensive overlap in the genetic basis between common mental disorders, functional (15), and structural connectivity of the brain (16,17). Thus, looking into the connectome for endophenotypes that link genetic risk with fear expression is a promising endeavor. Despite decades of research, well-studied brain regions still have not been fully characterized in their specific roles for different phases of fear memory processing. Besides, only a few regions have attracted the attention of researchers disproportionately, while others remain understudied. Specifically, the amygdala, the prefrontal cortex, especially its ventromedial subdivision (vmPFC) in humans, and the hippocampus have been established as the core components of the fear extinction network (18). However, a wealth of evidence propose the dorsal anterior cingulate (dACC) as a generator of homeostatic autonomic and behavioral responses according to the current interoceptive and emotional state represented therein (19). Furthermore, the cerebellum’s role on associative fear memory formation has been well characterized (20), but its involvement in fear extinction remained until very recently largely ignored (21). Similarly, the notion of the vmPFC being instrumental for successful extinction of conditioned fear is widespread (22) but that of its contributions to fear learning is not (23). In sum, recent work suggests that the core network should be expanded to include other critical regions due to their consistent involvement in both fear memory formation and extinction (19,24).

Here, we carried out exploratory mediation analysis to test the hypothesis that individual differences in PGS, as well as *DCC*- and learning ePGS, predict differences in SCRs of fear acquisition and extinction, mediated by structural and functional connections within the human fear network.

## Methods

This is a pre-registered work (https://osf.io/m34rd). See S*upplements* for any deviation from the pre-registration.

### Sample

Sample data stems from a German multisite collaborative research center focused on the study of extinction learning (SFB1280; https://sfb1280.ruhr-uni-bochum.de/). This study was conducted in accordance with the Declaration of Helsinki, and each study protocol was approved by the local ethics review board of the respective study site (see *Supplements*). For the present study, only participants with available DNA, skin conductance, resting state functional- and diffusion-weighted magnetic resonance imaging (rs-fMRI and DW-MRI, respectively) data of sufficient quality were considered. These criteria reduced the total sample size to 153 (68 men) with data available for analysis. The pooled age range was 1836 years with an average of 23.55 years and a standard deviation from the mean of 3.55 years.

### Polygenic Scores

We calculated conventional PGS by applying publicly available summary statistics of GWAS on major depressive disorder (25) (MDD), post-traumatic stress disorder (26) (PTSD), anxiety disorders (27) (ANX), neuroticism (28) (NEURO), as well as cross-disorder risk (29) (CROSS) on the present sample’s genotype data (for details on sampling, genotyping, preprocessing, and imputation, see *Supplements*). Moreover, we included GWAS for verbal short-term memory (STM) and verbal learning (VL) (30) on the premise that genetic load for fundamental cognitive capabilities, such as memory, might affect neural system communication, since the integrity of these capabilities might affect associative memories (31). PGS were calculated following guidelines recommended for ‘LDPred-2 auto’ (32,33).

### Trait-specific and Cross-trait PGS

It has been convincingly shown that mental disorders share genetic risk factors (7,34). Thus, we set out to explore the impact of pleiotropic covariation on PGS performance by adjusting GWAS SNP-effects of each trait according to genetic correlations to others. LD Score Regression (35) v1.0.1 was used to identify significant FDR-corrected genetic correlations. Next, we applied multi-trait conditional and joint analysis (36) (mtCOJO) to extract disorder specific SNP-effects, conditioned simultaneously on all other covarying traits. The method is a summary statistics-based approach that uses Generalized Summary-data-based Mendelian Randomization to test for a putative causal association between two phenotypes. The output of this procedure is a new set of summary statistics which were then used to construct ‘conditioned PGS’ (PGSc), as described in the previous section. Conversely, we used genomic structural equation modelling (37) (genomic SEM) to also capture the overarching genetic effect across traits in common latent factors. Accordingly, we sought to find a structural model that best explains genetic variance inferred from the summary statistics by means of exploratory (EFA) and confirmatory factor analyses (CFA) on the genetic covariance- and sampling covariance matrix. Next, the best-fit model was expanded to estimate the effect of any given SNP on the identified latent genetic factors. SNPs in LD (*r^2^* > 0.2) with genome-wide significant SNPs, with *p* < 0.5*10^-8^ for genomic SEM’s *Q* statistic (37), 0.4 > MAF > 0.1, or with negative estimated variance were excluded from the resulting cross-trait summary statistics. The remaining estimates were used for latent-factor PGS construction with LDPred-2 auto.

### Expression-based Polygenic Scores

GTEx gene read counts of 56 000 transcripts for the anterior cingulate cortex, frontal cortex, amygdala, hippocampus, and cerebellum served as the basis expression data for all ePGS. We used the R package GWENA (38) v1.6.0 with standard settings to construct weighted and unsigned co-expression networks and to identify modules of highly co-expressed genes on the preprocessed expression data. For each tissue, we curated a list with all genes within the module where *DCC* was allocated. Alternatively, we carried out over-representation analysis to identify modules that were significantly enriched in gene ontology (GO) terms with evidence of biological relevance for associative learning and listed the genes therein (see *Supplements*). Next, all cis-expression quantitative trait loci, and their respective effect on gene expression among all genes on the list formed the basis for ePGS construction in the final step. After clumping to account for LD (500kb window, R^2^<0.2), the overlap in remaining SNPs between GTEx and the present sample constituted the final set of variants to construct ePGS. The number of alternative alleles weighted by their gene-SNP association coefficient summed over all SNPs defined the individual measure of co-expressed *DCC*/learning network activity in a given brain region per participant. The final score used in mediation analyses was the sum of pairs of ePGS. Only ePGS from different regions of interest (ROIs) were summed together, yielding ten *DCC*-ePGS and ten learning-ePGS per subject.

### Structural and Functional Connectivity of Imaging data

Information on acquisition, preprocessing, and additional analytical decisions concerning imaging data is provided in the *Supplements*.

The partial correlation coefficient using a Ledoit-Wolf shrinkage parameter served as a measure of functional connectivity. It assesses more direct interactions between two brain regions and offers estimates with improved within-subject stability compared to marginal correlations (39), reflecting more trait-like aspects of brain organization. Having only five (bilateral) ROIs, we included the estimate of all ten possible ROI-pairings in downstream analyses.

Probabilistic tractography with FSL’s ProbtrackX (40) was performed to extract streamline counts as a measure of structural connectivity. Since the number of streamlines found between ROI pairs is not necessarily the same in opposite directions (e.g., the number of counts when streamlines start from the hippocampus and terminate in the amygdala is unequal to that of the same operation in the reversed direction) we opted for the average across both. As with functional connectivity, we obtained a bilateral estimate of counts for each of ten ROI pairs.

### Skin Conductance Responses

Skin conductance data was analysed with the Psychophysiological Statistical Parametric Mapping (PsPM v6.0.0) toolbox (41). Participant-specific learning measures were obtained by first fitting a 2^nd^ order polynomial regression on amplitude scores extracted from PsPM (see *Supplements*) as the dependent variable. Trial number, CS-type and their interaction were used as predictors. The model’s predicted value for each amplitude score was subtracted from the one succeeding it. The sum of all subtrahends represented the learning measure associated with a CS-type. Positive values indicate a net increase in SCR amplitudes with each additional trial, and a decrease in the case of negative scores. CS+ scores represent simple learning to conditioned fear stimuli, whereas CS-scores represent learning to conditioned safety stimuli. During acquisition, larger positive CS+ scores and negative CS-scores indicate more effective learning of conditioned fear and safety, respectively. During the extinction phase, fear is extinguished the more successfully the smaller CS+ scores are.

By subtracting CS+ from CS-scores we obtained differential learning scores. These scores reflect an individual’s ability to discriminate the CS+ from the CS-. The greater the score’s absolute value, the greater the CS-discrimination. Larger difference values imply better differential learning during acquisition. Conversely, differences closer to zero indicate better learning throughout extinction training, i.e., learning that CS+ stimuli no longer precede aversive outcomes and thus formerly threat-associated stimuli tend to elicit SCRs like safetyassociated cues over time.

### Statistical Analysis

All analyses were performed in R v4.2.2. We carried out exploratory mediation analysis by regularization using XMed (42) to identify a subset of variables with meaningful mediating effects. It is a process consisting of two phases that rely on elastic-net regression, which offers the advantage of reducing model complexity by eliminating irrelevant variables while also dealing with multicollinearity in the model. In the first phase, we set up a group-balanced 5-fold cross-validation scheme to assess model generalizability. However, results are likely to be unstable due to the modest sample size. To counteract this, we imposed additional restrictions on how to declare a true mediation effect as detected. Our approach consisted of repeating the above procedure 1000 times, reassigning observations to different folds each time. Out of all iterations, the median of the standardized mediation effect estimates was extracted alongside its corresponding bootstrap-based standard errors. If the absolute value of the estimate’s bounds was greater than 0.001, that value was considered evidence of a true mediation effect. Due to the conservative penalization introduced by the elastic-net, coefficient estimates for all predictors may be underestimated. Thus, in the second phase, mediation models were fitted again on the subset of meaningful mediators and predictors identified in the first phase but without penalization using lavaan’s SEM (43) with the robust maximum likelihood estimator and robust standard errors to correct for coefficient underestimation.

All PGS measures were treated as predictors. PGS, PGSc, and latent factor PGS were submitted to separate runs. All brain connectivity measures acted as mediators. The learning indices were regressed on the predictors and mediators separately for each experimental phase and learning score. Likewise, mediators of different modalities were submitted to separate mediation models. All models were controlled for genetic ancestry (first five genetic principal components), age, sex, group- and study of origin. The above procedure applies exclusively to PGS. ePGS, however, were not analysed with XMed since we do not assume activity levels of *DCC*/learning ePGS to be related to connectivity metrics other than to the brain regions they are constructed from. Instead, we ran mediation models on the PROCESS macro v4.2 (44) for R for each predictor-mediator pair separately.

## Results

### Genetic associations & SNP-effect modeling

Each trait showed substantial genetic correlations with several others (Figure S1). Correlations among disorder risk summary statistics were all positive and ranged from 0.67 to 0.84 (all *p_FDR_* < 0.001). The correlation between cognitive traits was estimated to be 0.89 (*p_FDR_* < 0.001). Disorder risk and cognitive traits correlated negatively, with NEURO and PTSD showing a correlation to VL and STM; MDD showed a moderate negative association with STM only (all *p_FDR_* < 0.01, Figure S1). Accordingly, we carried out mtCOJO on each trait by specifying the above found links as covariates to obtain predictors free of confounding, i.e., PGSc.

From genetic correlation analyses, the genetic covariance matrix was extracted and submitted to EFA to devise a genetic architecture for latent factors. A two-factor CFA model was specified based on the EFA results, where all disorders loaded onto one latent factor while both cognitive traits loaded onto another (henceforth ‘PSY-factor’ and ‘COG-factor’, respectively). Latent factors showed a moderate negative correlation (*r* = -0.21; Figure S2) The final configuration provided an excellent fit to the data (*χ*^2^(9) = 37.035; AIC = 63.035; CFI = 0.992; SRMR = 0.058) and a better fit than a single factor model (*χ*^2^(8) = 180.347; AIC = 204.347; CFI = 0.955; SRMR = 0.195). Consequently, a multivariate GWAS was run using this latent structure to estimate the effect of every SNP on each latent factor. After filtering, 360 681 PSY- and 357 172 COG-SNPs were available for ‘PSY-PGS’ and ‘COGPGS’ construction, respectively.

### Exploratory mediation analysis

Figure 1 displays the main results of the exploratory mediation analyses. All three types of PGS were predictive of at least one learning index type, during either fear acquisition or extinction training. Furthermore, all five fear network ROIs played a role in some capacity.

**Figure 1.**
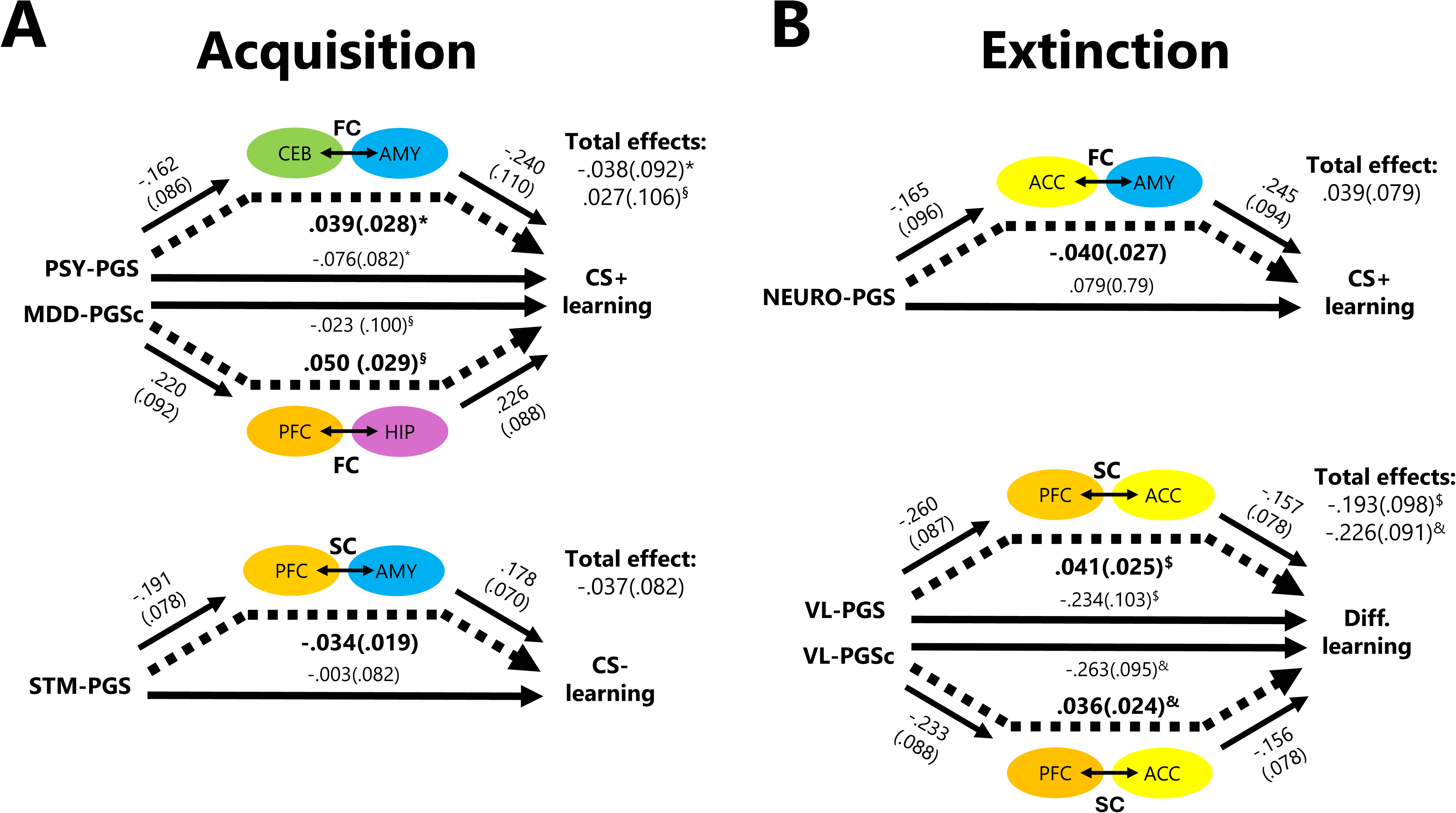
PGS prediction of acquisition and extinction of conditioned fear is mediated by structural and functional connectivity. Predictors and mediators that showed associations with CS+, CS- or differential learning during fear acquisition **(A)** and extinction training **(B)**. All coefficients displayed on top of arrows (standard errors in parentheses) are corrected for underestimation during elastic-net regression by re-running the mediation models without regularization and keeping only significant regressors identified during the first run (see *Methods*). Values in bold next to dotted arrows represent the indirect effects. The indirect effect is the result of multiplying coefficients together from paths pointing towards- and coming from mediators (short solid arrows). Each total effect is the sum of the estimates for the direct (values over long solid arrows) and indirect paths with the same superscript (*, §, $, &). Note that during fear acquisition no model predicted differential learning, and during fear extinction none predicted CS-learning. *AMY*, amygdala; *ACC*, dorsal anterior cingulate cortex; *CEB*, cerebellar nuclei; *FC*, functional connectivity; *HIP*, hippocampus; *MDD*, major depressive disorder; *NEURO*, neuroticism; *PFC*, ventromedial prefrontal cortex; *PGS*, polygenic score; *PGSc*, conditioned polygenic score; *PSY*, psychological latent factor; *SC*, structural connectivity; STM, (verbal) short-term memory; *VL*, verbal learning.

Conventional PGS construction of both STM and VL yielded predictions for CS- and differential learning, respectively. The former, however, only during fear acquisition and the latter only during extinction training. Structural connectivity between the vmPFC and the amygdala mediated the relationship between STM-PGS and CS-learning (Fig. 1A bottom). On the other hand, vmPFC-dACC structural connectivity mediated VL-PGS predictions of differential learning (Fig. 1B bottom). Regarding psychological traits, NEURO-PGS emerged as the sole relevant predictor of CS+ scores during extinction when jointly modelled with the functional connectivity between the dACC and the amygdala as an intermediary variable (Figure 1B top).

By constructing PGS conditional on genetically related traits, PGSc for MDD, i.e. free of genetic confounds, showed an association with differential learning during fear acquisition training that was mediated by vmPFC-hippocampal functional connectivity (Fig. 1A top). The connection found for VL was not altered after conditioning, except for minor numerical differences in the individual regression paths (Fig. 1B bottom).

Regarding latent factor-based PGS, PSY-PGS were predictive of CS+ learning during acquisition due to a mediation effect of cerebello-amygdalar functional connectivity (Fig. 1A top). No significant mediation involving the COG-factor was observed.

Since correlations among predictors ranged anywhere between faint and very strong (Figure S2), penalization dynamics could potentially fluctuate depending on the arrangement of variables submitted to elastic-net regressions. Thus, we further examined the effect of rerunning analyses with different predictor groupings. We included either only psychological, only cognitive, or each PGS individually and operated identically for PGSc or latent factor PGS. For PGSc analyses, we additionally specified models in which PGSc were tested alongside others for which there were no significant genetic correlations between their underlying traits, e.g., MDD-PGSc and STM-PGSc since MDD and STM summary statistics did not show a significant association (for an overview of model predictor combinations, see *Supplements*).

We found that the choice of predictor grouping did not change the selection of significant regressors. Averaging across different groupings, significant mediations using PSY-PGS were detected 81.5% of the time after 1000 5-fold cross validation iterations, providing the most stable prediction models overall, followed by MDD PGSc- (68.6%), NEURO-PGS (65.1%), VL-PGS- (59.3%), VL-PGSc- (56.0%), and STM-PGS models (Figure 2). STM-PGS, however, predicted CS-learning only when tested alongside all other PGS. On the contrary, VL-PGSc predicted differential extinction learning when no predictor other than themselves or STM-PGSc were submitted to the model. A similar case was found for CROSS- and VLPGS. Both predicted CS+ learning during fear acquisition and extinction training, respectively, but only when alone. Furthermore, these last two measures did not meet the criteria to pass onto phase two of exploratory mediation as the boundaries of their median effect estimate were not above threshold. Finally, we also tested all model arrangements as above but using a random control predictor and/or a control mediator in addition (see *Supplements*). There was no instance of a control variable showing evidence of stable associations to real data (for an exemplary visualization, see Figures S3 and S4).

**Figure 2.**
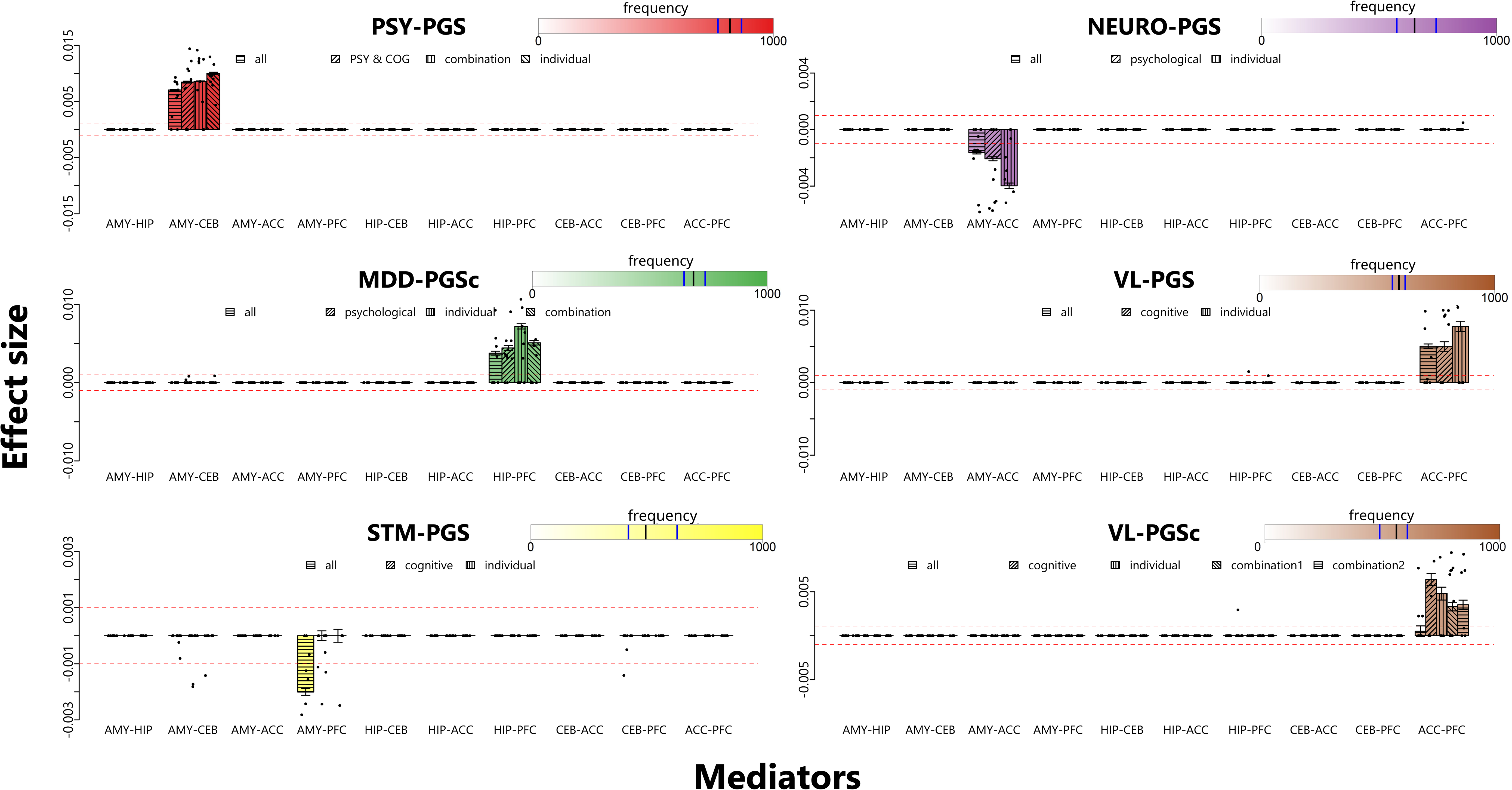
Mediation effects are robust to model specifications. Despite varying predictor combinations and sample compositions set up for analysis, the same PGS consistently showed significant effects specific to one mediator during phase 1 of exploratory mediation, leaving predictor and mediator selection for subsequent phase 2 models unchanged. Each panel corresponds to one path diagram in Figure 1. Y-axes display effect sizes, while mediators (i.e., ROI-pairs) are placed on the x-axis. Bars represent the estimated median effect found in a repeated cross-validation scheme with 1000 repeats. Each bar within each mediator represents the result of a different grouping of predictors (see main text and *Supplements*). Dots on bars are effect estimates from individual cross-validation iterations (only 10 are displayed for visualization purposes). Color intensity represents the frequency of effect detection. Black whiskers indicate the bootstrap-based standard error. Black and blue lines on the color scales represent the mean and range of detection frequency for significant mediators, respectively. To see which learning indices and connectivity modalities were included in the models, see Figure 1. Red dotted lines mark the 0.001 threshold beyond which effects are declared present in the data. ACC, dorsal anterior cingulate cortex; AMY, amygdala; CEB, cerebellar nuclei; COG, COG latent factor (see main text); HIP, hippocampus; MDD, major depressive disorder; PFC, ventromedial prefrontal cortex; PGS, polygenic score; PGSc, conditioned polygenic score; PSY, PSY latent factor (see main text); STM, (verbal) short-term memory; VL, verbal learning.

Taken together, PGS(c) showed robust specificity to one mediator associated with fear learning or extinction. Note, however, that for every unit increase of any PGS(c), the models predicted only marginal changes in learning indices.

### ePGS construction and mediation analyses

*DCC* was allocated to modules ranging in size from 111 to 301 co-expressed genes in all GTEx tissues of interest. Remarkably, most modules of either ePGS type were significantly and specifically enriched for several nervous system and neuronal learning GO pathways. To name a few examples, ‘learning and memory’, ‘neuronal synaptic plasticity’, ‘synapse organization’, ‘chemical synaptic transmission’, ‘establishment of localization’, and ‘structure development’ were recurring biological processes (all *p_corrected_* < 0.05). Moreover, cellular component terms specific to neurons such as ‘dendrites’, ‘postsynaptic membrane’, ‘dendritic spines’, ‘perikaryon’, ‘axon terminus’, among others, were overrepresented in these modules. For the cerebellar *DCC*-module, however, cellular compartments were cell-type non-specific and biological processes were not always focused on the nervous system. Table S2 summarizes WGCNA results, whereas Figures S5-S14 summarize overrepresentation analyses performed on each module selected for ePGS construction.

In sum, ePGS were based on SNPs affecting expression of gene networks with affinity for neuronal structure, organization and synaptic plasticity. Regardless, no significant mediation was found (all *p* > 0.05). It should be noted, however, that some direct paths, frequently those involving cerebello-hippocampal and amygdalo-hippocampal ePGS, showed an association with various learning indices at a nominal *p*-value level, but not after correcting for multiple comparisons (all *p_corrected_* > 0.05).

## Discussion

Susceptibility to fear learning and extinction is a highly heterogeneous, yet heritable trait (45,46). Here, we show that genetic disposition for mental disorders in a non-clinical population as well as genetic indices of cognitive capabilities are (indirectly) associated with individual variation in fear and extinction learning. As discussed below, constructing PGS while accounting for genetic interdependencies among traits improved prediction in some instances. Our results suggest an involvement of all fear network regions in at least one pathway connecting genetics and fear learning. Both functional and structural connectivity featured during fear acquisition and extinction training. Simple learning of the conditioned fear stimulus (CS+) was most reliably predicted via functional connectivity, whereas differences in safety cue (CS-) and differential learning (CS+ vs. CS-) were mediated by structural connection strength.

Several PGS implementations have shown the benefit of modelling pleiotropic associations among traits to improve prediction at the phenotype level (e.g., SNP effects of schizophrenia and bipolar disorder jointly modeled to predict the corresponding case-control status (47)). The reverse operation, i.e., estimation of trait-specific SNP effects conditioned on all others, leads to similar results (48). Here we show that the predictive power, occluded in marginal GWAS effect estimates, can be unveiled by leveraging pleiotropy. This was showcased by PSY-PGS and MDD-PGSc. Predictions including PGS derived from this PSY-factor yielded the most robust effect detection overall. Specifically, PSY-PGS showed an association with the acquisition of conditioned fear, mediated by partial correlations of BOLD responses at rest between the amygdala and the cerebellum. Higher PSY-PGS were associated with net increases in CS+ responding over time mediated via amygdalo-cerebellar functional connectivity. This finding adds to the growing body of evidence that point to the cerebellum and amygdala interacting to foster emotional control (49). High genetic load for mental disorders may predispose individuals to develop fear responses to aversion-coupled stimuli more rapidly by altering functional communication between these areas.

Moreover, higher MDD-PGSc predicted higher CS+ responding when modeling vmPFC-hippocampal functional connectivity as an intermediary variable. The indirect effect was positive because this mediator was positively associated to both MDD-PGSc and CS+ learning. Recent perspectives propose that the vmPFC prepares individuals in the face of anticipated danger, such as during the acquisition of conditioned fear (23,50). Neural events in the hippocampus are believed to encode context-related information of new episodic memory traces (51). Together, the prefrontal-hippocampal circuitry computes stimulus value to estimate the degree of threat tied to a stimulus (52). A positive association between MDD PGSc and functional hippocampal-prefrontal covariation might appear surprising at first, given that past research has shown hypoconnectivity between these areas in depression (53,54) as well as in anxiety- and stress-related symptomatology (55,56). Importantly, these disorders are highly comorbid and share a substantial portion of genetic influences (57). Incongruency with past research might be thus attributable to the disentanglement of MDD SNP effects from those of comorbid traits carried out here.

Interestingly, genetic risk for neuroticism showed a relationship to CS+ learning during extinction training via dACC-AMY functional connectivity. Functional dACC connectivity with the amygdala supports successful updating of fear associations (58). A failure to extinguish such associations has been linked to compromised integrity and hyperactivity of the dACC (59) and to increased AMY-dACC connectivity after fear acquisition training (60). Our models predicting more positive CS+ learning during extinction (i.e., inefficient extinction learning) for individuals with higher functional connectivity at rest are thus well in line with past findings and suggest high trait connectivity as a vulnerability factor for successful extinction of aversive memories. Counterintuitively, individuals at greater genetic risk for neuroticism tended to have lower functional connectivity estimates, which in turn led to more efficient extinction. Research by Pace-Schott et al. (61) demonstrated that decreased connectivity between the amygdala and ACC indeed corresponded to higher neuroticism, implying a detriment in regulatory neural mechanisms of negative affect (ACC exerting top-down control over amygdala activity) in individuals with elevated neuroticism. In turn, this could provide an unexpected advantage of more efficient extinction learning, as our results suggest. Moreover, neuroticism showed large genetic correlations to all disorders and the largest loading to the PSY-factor in our analyses, suggesting that individual variation on the latent factor most closely relates to variation in neuroticism. It is reasonable to presume that due to the shared architecture of neuroticism with other disorders, this trait captures variation in imaging and physiological markers much like the combined genetic liability of internalizing disorders. This interpretation finds support in other investigations that position neuroticism as the binding common denominator among internalizing disorders at the phenotypic level since genetic factors between neuroticism and internalizing disorders overlap greatly, though not entirely (62,63).

Notably, both PGS for memory capabilities predicted fear learning. An increased genetic disposition for short-term memory was associated with a net tendency for SCRs to decrease after each additional CS-stimulus presentation during fear acquisition protocols, depending on the strength of structural connections between the vmPFC and the amygdala. Physiological responses to the CS-increased with greater vmPFC-AMY structural connections. Conversely, STM-PGS were inversely related to connectivity between these ROIs. During the acquisition of fear, inhibitory efferents from the amygdala repeal the otherwise suppressive effect of the vmPFC on the amygdala-orchestrated expression of fear to favor the formation of CS-US associations within its basolateral subnuclei. The amygdala then initiates the propagation of a cortex-wide signal of several neuromodulatory systems to increase arousal and attention towards cues that predict danger (64,65). More extensive structural connections might then correspond to stronger amygdalar inhibitory influences on prefrontal regulation of excitability and a more disproportionate autonomic response to stimuli not linked to an aversive outcome. Genetic variation that promotes better performance in short-term memory tasks might help mitigate the detrimental effect of such predisposition on safety learning. On the other hand, more efficient differential learning during extinction training was indirectly related to lower (conditioned) PGS for verbal learning since the mediation path involving dACC-vmPFC structural connectivity was composed of antagonistic effects. Specifically, we found that stronger structural connections between the dACC and the vmPFC predicted a decrease in CS+/CS-learning differences, i.e., better differential extinction learning. Past research has shown that white matter integrity in the cingulum, a major fiber passing through these areas and connecting both to subcortical structures like the amygdala and the hippocampus, is positively related to (early) extinction learning (66). Thus, a greater number of streamlines between these cortical areas might reflect a better preserved pathway that facilitates the exercise of their canonical functions during extinction in interaction with other brain regions: stimulus value appraisal and fear response inhibition (67). Higher VL-PGS(c), however, were related to lower structural connectivity, ultimately slowing down extinction learning. This is surprising given that positive associations between structural connectivity and individual differences in memory performance are ubiquitous in the brain (68), including the superior cingulate bundle (69). It should be noted that mediation effects for cognitive traits were the least robust and depended on the exact combination of predictors in the model. CS-learning was linked to STM-PGS when every other predictor was submitted for analysis simultaneously, but not when STM-PGS were tested alongside VL-PGS or alone. Similarly, VL-PGSc allowed for above threshold predictions but not when competing against all regressors. Caveats notwithstanding, these results find some support for the idea that (genetic disposition for) memory related higher-order cognitive faculties may aid the formation and modification of CS-US contingencies.

Concerning ePGS analyses, the identified gene network modules were finely tuned for associative learning pathways at the molecular and cellular level. Despite biological plausibility, ePGS showed no evidence of association to learning, directly or indirectly. Though some direct paths from ePGS to learning hinted at a covariation between both, these analyses did not survive multiple comparison corrections. Thus, null results suggest that while ePGS may be related to fear conditioning, connectivity of the fear circuitry may not be a relevant intermediate variable.

The focus of the present work was on brain connectivity at rest, as opposed to task-based, on the premise that genetic composition partially affects trait-like measures, e.g., BOLD signal fluctuations in resting-state networks (70–72). However, future studies could examine the effect of polygenic markers on neural activation *during* fear extinction experiments, given previously reported associations between genetic variability and in-task modulation (73,74). Moreover, other imaging-based endophenotypes are needed to further characterize the intermediary pathway between polygenic predictors and learning, as evidenced by non-zero direct effects that point at variance yet to be explained.

## Conclusions

In the present work we set out to identify genome-wide distributed contributions to fear expression and found that additive genetic load for mental disorders, neuroticism, and memory-related cognitive capabilities have a rather modest, yet non-negligible impact, on the connectivity of the human fear circuitry, which in turn relates to individual fear learning and extinction behavior over time. Further, we conclude that while genetic correlations among traits are pervasive, these can be harnessed to better portray the relationship between genetic variation, neural systems, and ultimately, behavior in health and disease.

## Data Availability

All data produced in the present study are available upon reasonable request to the authors.

https://doi.org/10.17605/OSF.IO/M34RD

## Acknowledgements

This study was supported by the Deutsche Forschungsgemeinschaft (DFG, German Research Foundation) – Project-ID 316803389 – SFB 1280.

JESP conducted all analyses and wrote the manuscript. In addition to conceiving and developing the project, RK reviewed and edited all drafts. CAG processed imaging and electrodermal data. TS, EG, CJM, OT, DT, SE, HE, HQ, OG, and NA facilitated raw data sharing. All authors provided valuable comments when preparing the present work.

All authors declare that all relevant ethical guidelines have been followed, all necessary ethics committee approvals have been obtained, all necessary participant consent has been obtained and the appropriate institutional forms archived.

All authors declare no conflict of interest.

