## Supplements for "Polygenic prediction of fear learning is mediated by brain connectivity"

### Supplementary Sections

#### Methods

##### Contributing SFB studies

In the following, brief descriptions of the experimental setup of all six studies (S1-S6) stemming from four working groups (A03, A05, A09, A12) that provided data for the present work can be found (see also Table S1). Some studies included further fear conditioning phases (e.g., habituation, renewal) that are not mentioned below since we did not examine them.

*A03, studies S1 and S2.* The differential fear learning paradigm involved the presentation of visual stimuli (neutral pictures of an office setting) of which some were paired with electrical stimulation on the skin. The stimuli and procedures were modified from Milad et al(1). Visual stimuli were presented using the Presentation software package (Neurobehavioral Systems, Berkeley, CA, USA) and MR compatible LCD-goggles (Visuastim Digital, Resonance Technology, Northridge, CA, USA). Electrical stimulation (1 ms pulses at 50 Hz for a duration of 100 ms) was applied using a constant voltage stimulator (STM2000, BIOPAC Systems, Goleta, CA, USA) along with two electrodes attached to the fingertips of the index and middle finger of the right hand. The intensity of electrical stimulation was adjusted for each participant individually prior to the fear acquisition phase. Electrical stimulation was administered at 30 V and raised in increments of 5 V until participants rated the sensation as very unpleasant but not painful.

Each trial started with the presentation of a white fixation cross on a black background for 6.8 - 9.5 seconds. Next, a context image showing an office room with a switched-off desk lamp was presented for 1 second. This was followed by the same image but with the desk lamp either emitting blue (CS+) or yellow light (CS-) for another 6 seconds. In the acquisition phase, the CS+ was paired with electrical stimulation in 62.5 % of trials. Electrical stimulation was administered 5.9 seconds after CS+ onset and co-terminated with CS+ offset. CS+ and CS- were presented 16 times each in pseudo-randomized order but distributed equally across both halves of the acquisition phase. The first two trials always involved one CS+ and one CS- presentation and so did the last two trials. The first and last CS+ presentations were always paired with electrical stimulation. The same type of CS was not presented more than twice in consecutive order. The extinction phase consisted of 8 CS and 8 CS- presentations without US pairings.

*A05, study S3.* The study used a differential fear conditioning paradigm based on Ernst et al(2)., using black-and-white geometric figures (a square and a diamond) of identical brightness as CS. During fear acquisition training, the CS+ was paired with an aversive US in 62.5% of trials, while the CS- was never followed by the US. The two CS figures were pseudo-randomly counterbalanced across participants. The experiment consisted of acquisition training (10 paired CS+/US trials, 6 CS+ only trials, 16 CS- only trials), and extinction training (16 CS+ only trials, 16 CS- only trials). Trial order was pseudorandomized, with specific constraints: the initial two and last trials of the acquisition training were paired CS+/US trials. Additionally, an equal number of events for each type were presented in both the first and second halves of each phase. The sequence of events remained consistent for all participants throughout acquisition and extinction. The electric shock was delivered via a constant current stimulator (DS7A, Digitimer Ltd., London, UK) to the left shin using a concentric bipolar surface electrode with a 6 mm diameter and central platinum pin (WASP electrode, Specialty Developments, Bexley, UK). The US comprised four consecutive 500 µs current pulses with a 33 ms interval, with intensity determined at the start of the experiment. Participants rated sensation intensity until it reached a score of 8 out of 9 on a Likert scale, corresponding to the subjective impression of “unpleasant but not painful”. Individual thresholds were increased by 20% to prevent habituation. Each trial involved an 8-second presentation of the CS. In reinforced trials, a 100-

millisecond US was presented after 7.9 seconds and ended at the same time as the CS. Intertrial intervals were randomized between 14.3 s and 17.9 s. Each part of the experiment was completed during a different session of fMRI data collection.

*A09, study S4*. Conditioned stimuli (CS) consisted of white geometrical shapes (rhomb, parallelogram and square) with similar luminescence on a black background. To ensure similar luminescence, all white geometrical shapes were designed to match in surface area between stimuli. Each trial had a total duration of 20s and consisted of a black screen presented for 0-2.5 s at the beginning followed by an 8s presentation of the CS and a 9.5-12 s inter-trial interval (see 3). The three shapes were randomly assigned to the three CS and balanced between groups. The US consisted of a 100 ms electrical stimulation applied to the fingertips of the participant's right index- and middle-finger via two 1 $cm^2$ electrodes using a constant voltage stimulator (STM200; BIOPAC systems, CA, USA). The stimulation level was adjusted individually to be unpleasant but not painful(3). The paradigm was presented via MR suitable LCD-goggles (Visuastim Digital, Resonance Technology Inc., Northridge, CA, USA) and realized in Matlab 2017a (Mathworks Inc., Sherborn, MA, USA).

Fear acquisition training on day one served to establish a CS-US association: two of the three CS (CS+G and CS+N) were immediately followed by the US in 5/8 trials whereas the CS- was never followed by the US (8 trials). Importantly, there was no difference in the fear acquisition protocol for CS+G and CS+N. Prior to fear acquisition training, participants were instructed to pay attention towards possible associations between the presentation of a geometrical shape and the electrical stimulation because they would be asked about them using a questionnaire afterwards. Participants were classified as contingency aware in case they were able to correctly identify the two CS+ (sometimes followed by an electrical stimulation) and the CS- (never followed by an electrical stimulation) after fear acquisition training(3,4). Before extinction training, the participants were informed that the associations learned on day one would not change over the course of the entire experiment to avoid expectancy of contingency reversal. Importantly, participants were not informed about the actual contingencies on any of the three experimental days. For more details from this study, see (3).

During extinction training on day two, the CS+N and the CS- were presented solely in their original size testing the effects of standard extinction training. The CS+G, however, was additionally to its original size presented in three smaller sizes (75%, 50% and 25% of the original size) to test for the effects of generalised extinction training. The three CS were presented eight times each during extinction training. Each size of the CS+G was presented two times to reach a total number of 8 extinction trials. Learning effects based on a higher number of presentations (e.g., 8 for each size of the CS+G) were avoided using this approach.

*A12, studies S5 and S6.* Both were two randomized-controlled studies conducted in independent samples of healthy volunteers. Both studies involved two consecutive days, with acquisition training on day 1 and extinction training accomplished 24 hours later. Participants were randomized to intravenous injection of lipopolysaccharide (LPS), as an established experimental model of acute systemic inflammation, or to an injection of saline as a placebo. Injections were accomplished two hours before either aquisition (study 1) or extinction training (study 2). Paradigms in both studies were identical, with visceral pain induced by pressure-controlled rectal distensions and equally unpleasant, aversive tones implemented as visceral unconditioned stimulus (US+vis) and auditory US (US+aud), respectively, during acquisition. Note that prior to acquisition training, US stimulation intensities were individually calibrated and matched for perceived unpleasantness within a predefined perceptual unpleasantness range of 60–80 mm on a 0–100 mm visual analogue scales (based on Koenen et al.(5)). As CS, three distinct visual symbols were paired with either US+vis (CS+US+vis), or US+aud (CS+US+aud), or were presented without US (CS-) during acquisition training. The reinforcement schedule was 75%, with 18 US (9 US+vis, 9 US+aud) and 36 CS presentations (3 out of 12 CS+ presentations were not followed by a US). All CS+ were presented 6–12 s before US (US durations: 14 seconds), with CS and US co-terminating. During extinction training, all CS were presented without US using the same pseudorandomized CS sequence as during acquisition training. In all phases, inter-stimulus intervals consisted of a black screen with a fixation cross with a duration of 8 s. Electrodermal activity was continuously recorded using an MRI-compatible system (Biopac Systems, Inc., Goleta, CA, USA; MP160). Only CS+ US+ vis and not CS+US+aud were included in the present work, and analyses included data from participants treated with LPS or placebo. For methodological details and previously published data from these studies, see (5,6).

#### DNA Sampling, Genotyping, Preprocessing, and Analysis

Exfoliated cells from the participants' oral mucosa were collected. After collection, the samples were transferred to the West German Biobank (WBE) at the University Hospital Essen and stored at -80°C until further analysis. The QIAamp DNA mini-Kit (Qiagen GmbH, Hilden, Germany) and Illumina's Infinium Global Screening Array 1.0. with MDD and Psych content (Illumina, San Diego, CA, USA) were used for DNA isolation and genotyping, respectively. Genotyped SNPs with a minor allele frequency (MAF) lower than 5%, deviating from Hardy-Weinberg equilibrium (HWE) with $p<1x10^{-6}$ and missingness higher than 2%, were excluded. Participants showing sex-mismatch, a SNP-missingness rate higher than 2% and heterozygosity rate higher than |0.2| were excluded. Furthermore, genetic relatedness filtering was carried out on a SNP-subset showing high genotyping quality (HWE $p>0.02$, MAF>0.2, SNP-missingness of 0%) and pruned for linkage disequilibrium ($r^2=0.1$). Pairs of cryptically related subjects with a pi hat value > 0.2 was applied to exclude subjects at random. Principal components (PCs) were computed to control for population stratification. Any participant deviating by more than |4.0| standard deviations from the mean on at least one of the first 20 PCs were also excluded. All filtering steps were performed using PLINK 1.9(7). The filtered genotype data was imputed using the European population of the Haplotype Reference Consortium (r.1.1 2016; GRCh37hg19). We chose Eagle 2.4 for phasing and Minimac4 for imputation. After imputation, we reduced the SNP-pool to well-imputed and genotyped common variants ($R^2 > 0.9$ and MAF>0.05), leaving 6,180,980 SNPs remaining for further analysis.

#### Polygenic Scores

We checked and corrected for common misspecifications in GWAS summary statistics and standardized all files using Bioconductor's MungeSumstats(8) v1.4.5.

#### Pleiotropy-informed Trait-specific and Cross-trait PGS

Since ANX, VL, and STM summary statistics lacked a sufficient number of independent genome-wide significant SNPs for GSMR/mtCOJO at the typical threshold of $p < 5x10^{-8}$ after clumping (>1MB apart or $r^2 < 0.05$), we relaxed the significance threshold to $p < 5x10^{-6}$.

We left summary statistics of cross-disorder risk out of mtCOJO- and genomicSEM analyses. Cross-disorder summary statistics are themselves a product of a meta-analysis of several common mental disorders(9). Conditioning on other traits SNPs would render the resulting SNP effects uninterpretable. Conversely, including cross-trait effects to find cross-trait latent factors would be redundant.

#### Expression-based Polygenic Risk Scores

*Preprocessing of expression data.* Having ranked all transcript by their mean expression, we kept only the transcripts with the highest mean expression whenever multiple transcripts corresponded to a single gene and removed the bottom 50% to reduce noise. A variance stabilizing transformation was then performed to account for mean-variance dependencies. Next, each gene's variance was modeled as an inverse gamma distribution and further filtered based on a p-value cut-off of 0.1 with CEMiTool's 'select_genes' function(10). Lastly, a partial PC-correction as per Parsana et al(11). was carried out on the quantile-normalized data due to known batch-effects in GTEx RNA-seq data(12). The DCC gene was spared from filtering to ensure its inclusion in the next steps.

*Learning ePGS.* To select an appropriate co-expression module, we inspected the GO terms for each domain, i.e., biological process, molecular function, and cellular compartment, for which each module was significantly enriched, and picked the one with the most terms across domains relevant for associative learning such as synaptic plasticity and long-term potentiation. This process was done for each of the five GTEx brain tissues that correspond to the ROIs in our sample. Since most modules comprised several hundreds of genes, the number of enriched terms was copious, ranging from a few dozens to a couple hundreds. To reduce the manual inspections to a manageable amount, we opted for Bioconductor's 'simplifyEnrichment' v1.6.1. for R(13). SimplifyEnrichment summarizes all enriched terms by generating semantic similarity matrices to cluster terms together into groups and assigns cluster labels that are maximally representative of all constituent terms. The main output are word cloud diagrams alongside the cluster-wise rearranged similarity matrix. For each GO domain in each module belonging to each of five brain tissues we inspected the cluster labels. Supplementary figures S7-S16 display labels of the biological processes for the modules that seemed most appropriate for ePGS construction to our judgement. Some examples of recurring cluster labels found for biological processes terms are ''signaling", "learning", "projection", "transsynaptic", "behavior", "morphogenesis". In the case of molecular functions, "protein-coupled", "transmembrane activity", "postsynaptic signaling", "receptor" and cellular compartments, "synapse", "postsynaptic membrane", "dendrite", "axon", and "neuron" appeared multiple times.

#### Acquisition, Preprocessing, and Analysis of Imaging data

*Image acquisition.* MRI images were acquired using three different 3T MRI scanners. In the following, we provide a list of parameters for anatomical, functional, and diffusion imaging sequences for each site. In parentheses, we indicate the respective scanner and contributing study (see Table S1).

Parameters for high-resolution T1-weighted (MP-RAGE) anatomical images: TR = 8 ms, TE = 4 ms, flip angle = 8°, voxel size = 1 x 1 x 1 $mm^3$, FOV = 24 x 24 cm (Phillips Achieva; S1,S2,S4); TR = 2.53 ms, TE = 2 ms, flip angle = 7°, voxel size = 1 x 1 x 1 $mm^3$, FOV = 19.2 x 25.6 cm (Siemens MAGNETOM Vida; S3); TR = 1.77 ms, TE = 3 ms, flip angle = 8°, voxel size = 1 x 1 x 1 $mm^3$, FOV = 19.2 x 25.6 cm (Siemens Skyra; S5,S6).

Parameters for rs-fMRI (whole-brain T2*-weighted images using a gradient echo, echo-planar imaging (EPI) sequence): TR = 2.5 s, TE = 30 ms, flip angle = 90°, voxel size = 3 x 3 x 3 $mm^3$, FOV = 24 x 24 cm, 80 x 80 voxels, number of slices = 47, number of volumes = 190 (Phillips Achieva; S1,S2,S4); TR = 1.43 s, TE = 30 ms, acceleration factor = 2, flip angle = 69°, voxel size = 3 x 3 x 3 $mm^3$, FOV = 24 x 24 cm, 80 x 80 voxels, number of slices = 48, number of volumes = 190 (Siemens MAGNETOM Vida; *S3*); TR = 2.5 s, TE = 30 ms, flip angle = 90°, voxel size = 3 x 3 x 3 $mm^3$, FOV = 28 x 31 cm, 92 x 92 voxels, number of slices = 46, number of volumes = 192 (Siemens Skyra; S5,S6).

Parameters for DW-MRI: TR = 9.5 s, TE = 88 ms, flip angle = 90°, voxel size = 2 x 2 x 2 $mm^3$, FOV = 24 x 24 cm, 112 x 112 voxels, number of slices = 60, number of directions = 60 (b = 1000 $s/mm^2$) (Phillips Achieva; S1,S2,S4); TR = 5.5 s, TE = 114 ms, flip angle = 90°, voxel size = 1.6 x 1.6 x 1.6 $mm^3$, FOV = 24 x 24 cm, 132 x 128 voxels, number of slices = 60, number of directions = 60 (b = 1000 $s/mm^2$) (Siemens MAGNETOM Vida; study of group A05); TR = 10.2 s, TE = 87 ms, flip angle = 90°, voxel size = 2 x 2 x 2 $mm^3$, FOV = 60 x 60 cm, 120 x 120 voxels, number of slices = 70, number of directions = 60 (b = 1000 $s/mm^2$) (Siemens Skyra; S5,S6).

*ROI extraction.* Two different parcellation maps from Freesurfer (Desikan-Killiany and Destrieux; version 6) were used to extract the following ROIs: amygdala, hippocampus, ventromedial prefrontal cortex, dorsal anterior cingulate cortex, and cerebellar nuclei, for both the left and right hemispheres. In the cerebellum, only cerebellar nuclei ROIs were extracted since they are the sole output structure of the cerebellum. For this, we employed the SUIT pipeline (https://www.diedrichsenlab.org/imaging/suit.htm). After aligning the T1w image of each subject to the ACPC, the cerebellum was cropped from the rest of the brain and normalized using DARTEL for the purpose of matching it to the SUIT atlas in MNI space. We then applied an inverse normalization to reslice the SUIT atlas into the functional space of each participant. Finally, the cerebellar interposed, dentate and fastigial nuclei were extracted and merged into one single ROI.

Amygdala and hippocampus were taken from the automatic volumetric segmentation of the subcortical regions. We extracted the labels from the Desikan-Killiany atlas “medial-orbito frontal” and “caudal anterior cingulate”, which correspond to the vmPFC and dACC, respectively.

All ROIs were resampled into functional (rs-fMRI) and Freesurfer space. For functional connectivity, functional space was used. For structural connectivity, all ROIs were kept in Freesurfer’s native space wo we could take advantage of surface files (e.g., pial surface). In addition, the vmPFC and dACC volumetric ROIs were also converted into surfaces. These surface ROIs were only used for computing streamlines during probabilistic tractography.

We also extracted the bilateral thalamus using Freesurfer’s automatic segmentation. The thalamus of the contralateral hemisphere with respect to seed/target cerebellar ROIs was used as a waypoint to guide probabilistic tractography (see main text).

*Preprocessing of rs-fMRI.* After removing the first two volumes from the raw data, we performed motion- and slice timing correction, coregistration to the T1w image, resampling of the BOLD time-series onto native space using fmriprep v.20.1.1, and denoising according to Satterthwaite’s method (14).

*Preprocessing of DW-MRI.* MRTrix3’s dwidenoise and mrdegibbs functions(15) were used to denoise and remove Gibbs ringing artifacts from raw DW-MRI data, respectively. Susceptibility-induced distortions, eddy-current- and motion correction was performed in FSL v5.0.0(16). Next, we fitted a diffusion tensor model at each voxel, which returned the first three eigenvectors and corresponding eigenvalues, as well as a fractional anisotropy (FA) map. We used the FA map to create all the necessary transformation matrices from diffusion to anatomical- and freesurfer spaces. All transformation matrices were computed using boundary-based registration. Next, we modelled crossing fibres and created distributions on diffusion parameters at each voxel with FSL’s BEDPOSTX.

*Structural connectivity.* Following suggestions by Glasser et al.(17), we seeded from surfaces instead of volumetric ROIs to track fibers to and from cortical regions. For the tracking of fibers from cortical regions we used surfaces instead of volumetric ROIs. For subcortical regions and cerebellar nuclei, however, we used volumetric ROIs due to a tendency of these regions to contain a high degree of anisotropy. To guide tractography, we used the pial mask as a stopping mask, which prevented tracts from crossing the grey/white-matter interface. Furthermore, we excluded streamlines that did not stop by the target region. Finally, we also constrained tracts to bypass any regions containing non-white matter voxels (e.g., CSF, skull). We restricted the movement of cerebellar tracts by defining two stop masks (the pial surface ipsilateral to the cerebellar nuclei seed, and the cerebellum hemisphere) and by using the thalamus as a first waypoint, such that only tracts that initially run via the thalamus before reaching the target ROI are considered valid streamlines. The rationale for the latter is that most cerebellar nuclei project to different regions of the thalamus.

#### Acquisition, Preprocessing, and Analysis of Skin Conductance Data

Skin conductance responses (SCRs) were measured with Ag/AgCl electrodes filled with an isotonic (0.05 NaCl) electrolyte medium attached to the hypothenar of one hand of each participant across all studies considered.

The raw data was trimmed to the task duration and filtered using a median filter with varying amount of time points, such that an optimal number could be found that best reduced gradient artefacts. Alongside visual inspection and manual pruning, residual artifacts were detected using a script based on an automated quality assessment procedure(18).

After preprocessing, SCRs in the artifact-free data for all but two studies were modelled using Dynamic Causal Modelling (DCM) since this approach is most appropriate when the CS presentation is longer than 3-4 seconds. For both acquisition and extinction training, we modelled the phase context (fixed event) occurred during CS onset (fixed event), CS interval (flexible event) and US (fixed event). Note that PsPM assumes that the event sequence is the same for all trials, so in the case of non-reinforced trials, i.e., CS+ stimuli without subsequent US presentation (CS+US-), we modelled a non-US by adding an event of the same duration as the actual US in the place where the shock would have occurred during reinforced trials. Owing to the long US presentation time (14 seconds) of studies S5 and S6, however, we analysed spontaneous fluctuations (SF) in skin conductance for acquisition-phase datasets. The number of SFs was estimated using a non-linear model, which is believed to be more sensitive to tonic arousal(19). We opted to use the duration of the experiment to calculate SFs and subsequently split the estimated SCR amplitudes by their onset into CS and US.

After computing the amplitudes on a trial-by-trial basis for all studies, we summed these amplitudes and divided them by the duration of the corresponding CS trial, thus providing us with a representative amplitude score per trial. We proceeded as described for each trial, stimulus type, and experimental phase of each subject. Some studies contained less than four non-reinforced CSs in their experimental design, which did not allow for an accurate assessment of learning across time. In those cases, we combined CS+US+ and CS+US- trials in our modelling.

To discard the possibility that the DCM-estimated SCRs to the CS could have been confounded by the response to the US, we applied two additional steps. First, we gradually decreased the time interval between CS and US onset to the point that there would be no differences between CS+US+ and CS+US-. Second, given that the sudomotor response has roughly the shape of a Gaussian curve, for each trial we ensured that the onset of the Gaussian bump plus at least 2 standard deviations did not overlap with the onset of the US. Indeed, the difference between CS+ and CS+US- was not significant ($p > .10$), suggesting that our method appropriately disambiguated both types of response.

#### Statistical Analyses

Both the aversive unconditioned stimuli used to establish Pavlovian learning, and the conditioned stimuli differed across studies in regards of presentation frequency, conditioned stimulus type, unconditioned stimulus onset after conditioned stimulus presentation. An effort to account for this variability across studies is made, nevertheless, by coding the study and research group of origin as covariates as well as z-scoring all variables withing each study/group (but see *Pre-registration Deviations* further below).

Exploratory mediation analysis does not rely on p-values to determine the statistical significance of a mediation effect. Hence, it does not require a standard correction procedure for multiple comparisons. Whenever relying on the frequentist approach, however, the α-level was set to 0.05. We chose the false discovery rate (FDR) approach to handle multiple testing.

*Control analyses.* XMed is known for trading off a reduction in false negatives for an increase in false positives, as compared to conventional bootstrapping methods (20,21). That is, XMed tends to minimize the chance of not detecting true mediation effects at the cost of also detecting (a smaller proportion of) spurious effects from noise variables. This behavior is accentuated the smaller the sample size is and the smaller the mediation effects are. To safeguard against these biases, we performed control analyses in which one random variable was included as a predictor and another one as a mediator. This way, we can determine if the rate of effect detection for variables of interest is different from that of noise. Observations for the control variables were sampled from a random normal distribution. By chance, randomly sampled variables may be correlated to real data. Thus, for control analyses we performed the same as outlined above one hundred times, each time creating a random variable anew and averaging all mediation effects over iterations since the average likely nullifies chance correlations of individual iterations.

#### Results

##### Second Phase Exploratory Mediation Analysis

Due to singularity issues, fitting SEM models during the second phase of exploratory mediation was prohibitive unless dummy variables encoding the studies for which each participant was recruited were dropped. Accordingly, we estimated second phase models without these covariates.

##### Regressor Groupings

As described in the main text, we examined the impact of different regressor groupings. One possibility is to group mtCOJO-adjusted PGS according to genetic correlations among unadjusted summary statistics. Specifically, summary statistics of outcome traits were conditioned only on exposure traits for which there were significant genetic correlations (Figure S2). PGSc of uncorrelated traits and CROSS-PGS could be submitted as competing predictors of exploratory mediation analyses as their polygenic signal can be assumed to be largely non-overlapping. In the following, a list of predictors in each tested grouping that results from this rationale is provided. Combinations referenced in the plot hatches of Figure 2 are indicated in parentheses.

Grouping 1: ANX, VL, CROSS (lower right panel “combination2”)

Grouping 2: ANX, STM, CROSS

Grouping 3: MDD, VL, CROSS (lower right panel, “combination1”; middle left panel, “combination”)

Grouping 4: NEURO, CROSS

Grouping 5: PTSD, CROSS

#### Pre-registration Deviations

In the pre-registration, we failed to report that analyses including ePGS would not be run using exploratory mediation. Exploratory mediation is only admissible when dealing with multiple mediators. In the manuscript we clarify that in the case of ePGS we consider only single-mediator regression models to be a sensible test of our hypotheses.

We originally stated our intentions to evaluate inferential statistics on the second phase of the exploratory mediation. This is however not necessary since the generalizability of mediation effects is already addressed in the first phase. The second phase’s sole purpose is to provide unbiased regression coefficient estimates on the subset of identified meaningful mediators during phase 1, ignoring *p*-values altogether.

We failed to z-transform each participant’s data to their group mean and standard deviation *during* the cross-validation procedure since XMed does not allow such operations. To assess the impact of the deviation from this common practice in predictive modeling we mimicked XMed using the caret package, which does allow for this operation. We successfully replicated our results, indicating that the effect of within cross-validation z-scoring in this case was inconsequential.

Initially, we set out to use prostate cancer (PrCa) summary statistics (22) to generate a control PGS for which we did not expect any association with other measures. However, this trait is likely not a reasonable choice since it has been shown that there are differences in structural connectivity of the brain and cognition between healthy controls and untreated PrCa patients (23). Furthermore, there is evidence of genetic correlations and PGS correlations between diseases and traits beyond theoretical expectations(24,25), making the search for a suitable alternative control trait especially difficult. To make sure that we test a measure truly independent from genetic, imaging and electrodermal data we opted for the generation of noise variables, as described in the main text.

### Supplementary Tables

**Table S1**

*Contributing Research Groups and Sample Composition*

| SFB1280 Project Number | Studies | Number of Participants |
|---|---|---|
| A03 | S1, S2 | 76 |
| A05 | S,3 | 27 |
| A09 | S4 | 12 |
| A12 | S5, S6 | 38 |

*Note.* SFB1280 is a multisite collaborative research center focused on the study of extinction learning. An extensive description of the research collaboration as a whole and of each research group can be found at https://sfb1280.ruhr-uni-bochum.de/

**Table S2**

*Gene co-expression networks*

| | AMY | ACC | CEB | FC | HIP |
|---|---|---|---|---|---|
| Genes analyzed | 1577/1576 | 1538/1537 | 1417/1416 | 1528/1527 | 1648/1647 |
| WGCNA Soft-thresholding Power | 6/4 | 3/4 | 3/3 | 4/4 | 4/3 |
| WGCNA Scale-Free Topology Fit ($R^2$) | 0.900/0.851 | 0.848/0.779 | 0.967/0.950 | 0.813/0.860 | 0.903/0.860 |
| Modules identified | 12/11 | 14/8 | 12/12 | 13/11 | 8/13 |
| Genes not in Modules | 258/55 | 4/151 | 182/137 | 146/148 | 296/6 |
| Selected Module Size (Module No.) | 197(2)/397(1) | 169(2)/416(1) | 212(1)/155(3) | 111(6)/213(1) | 301(2)/324(1) |
| ePGS SNPs available | 506/23911 | 699/29608 | 91/5554 | 655/7132 | 902/14765 |

*Note.* Networks are based on gene read count data from GTEx v7. Results for DCC and learning gene networks are to the left and to the right of the slash, respectively. AMY, amygdala; ACC, anterior cingulate cortex; CEB, cerebellum; ePGS, expression-based polygenic risk score; FC, frontal cortex; HIP, hippocampus; SNP, single-nucleotide polymorphism; WGCNA, weighted gene co-expression network analysis.

#### Supplementary Figures

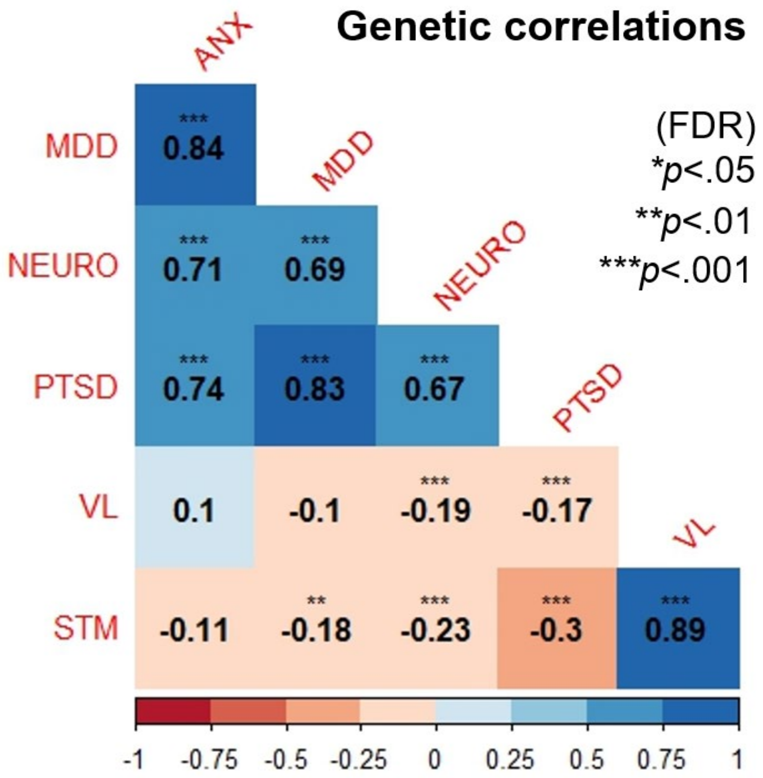

***Figure S1*. Genetic correlations among studied traits.** ANX, anxiety disorders; MDD, major depressive disorder; NEURO, neuroticism; PTSD, post-traumatic stress disorder; VL, verbal learning; STM, (verbal) short-term memory; FDR, false discovery rate.

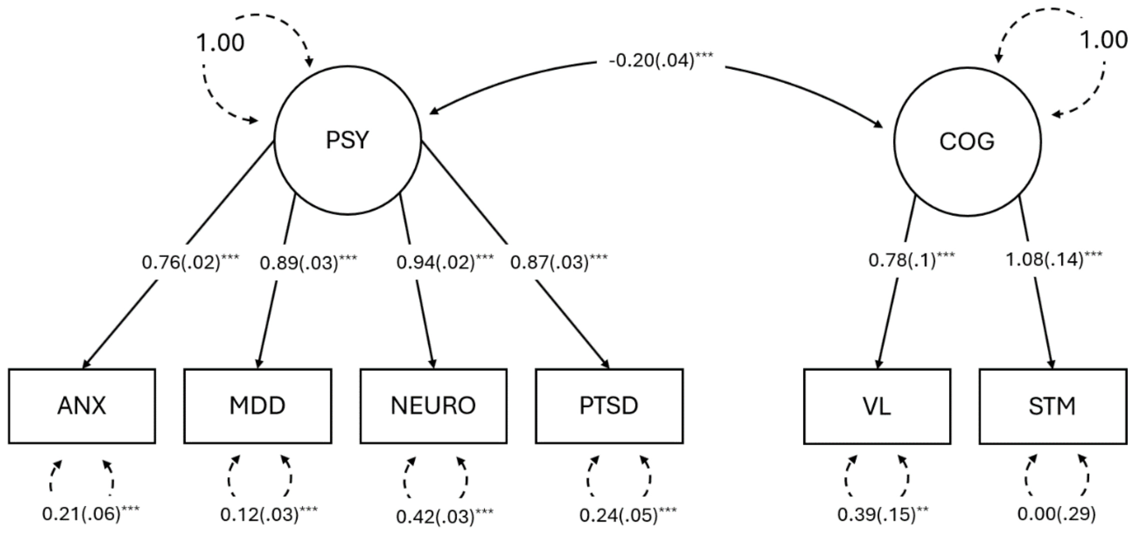

***Figure S2.*** **Confirmatory factor analysis without SNP-effects.** A two-factor solution, where both latent factors correlate, provided the basis for the multivariate GWAS. Standardized factor loadings and correlation between factors are indicated by values on top of solid lines. Estimates of residual genetic variance are indicated by values on top of dotted lines. Values between parentheses represent standard errors. Note that the factor loading for STM exceeds 1, which implies negative residual variance. Residual variance for this trait was therefore explicitly specified to be greater than 0. ANX, anxiety disorders; MDD, major depressive disorder; NEURO, neuroticism; PTSD, post-traumatic stress disorder; VL, verbal learning; STM, (verbal) short-term memory; ***p*<0.01, ****p*<0.001.

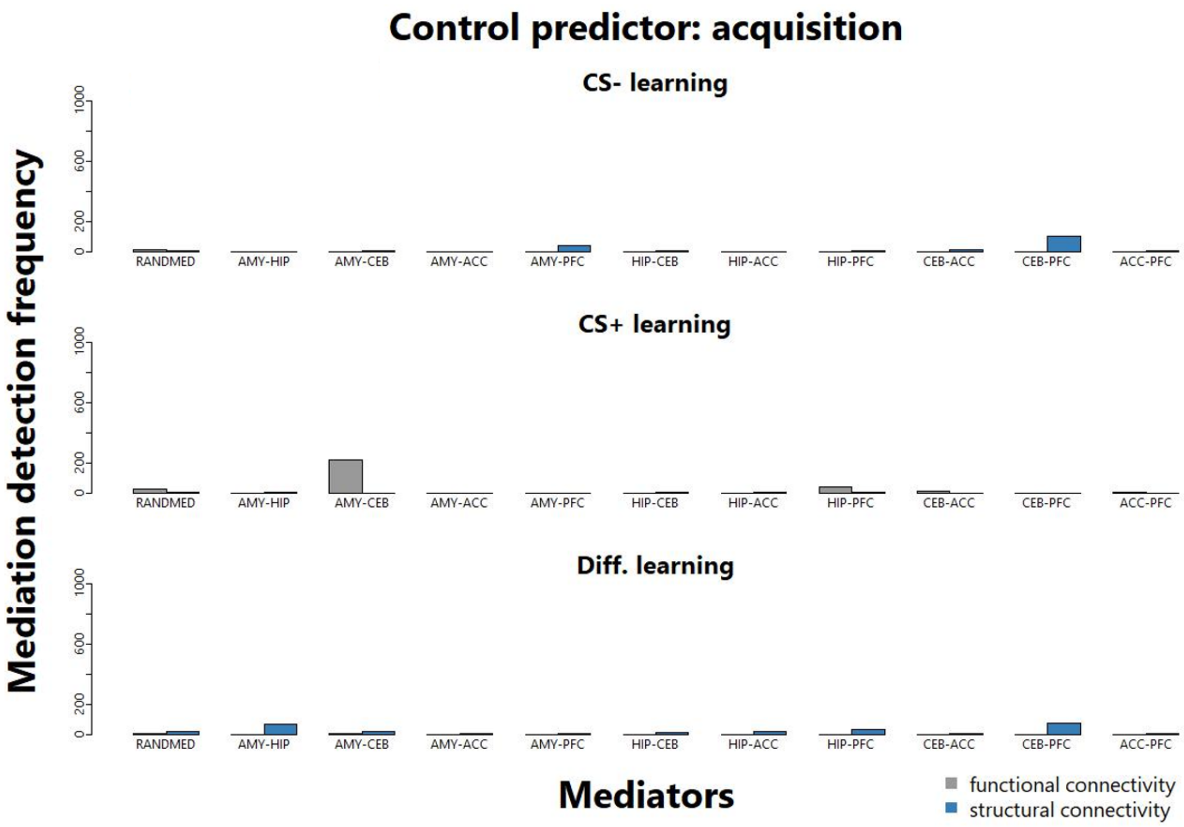

***Figure S3.*** **Mediation detection frequency of the control analysis for the acquisition of fear.** Exploratory mediation was carried out using a predictor and a mediator drawn from normally distributed random numbers to evaluate the rate of false positive detections in our dataset. Bar height represents the number of times a mediation effect for a specific ROI-pair surpassed the detection threshold when regressing each learning index onto the control predictor in a repeated cross-validation scheme with 1000 repeats. Results for functional and structural connectivity mediators are indicated by grey and blue bars, respectively. ACC, dorsal anterior cingulate cortex; AMY, amygdala; CEB, cerebellar nuclei; HIP, hippocampus; PFC, ventromedial prefrontal cortex.

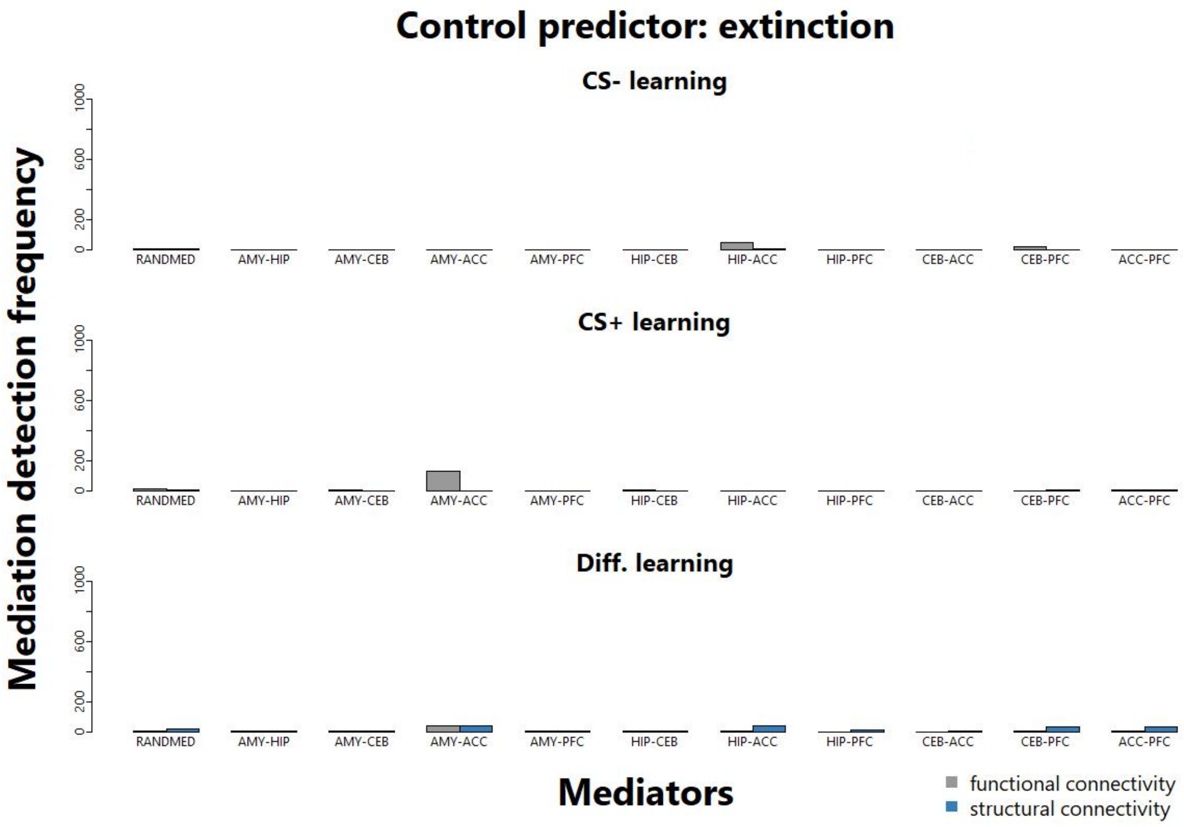

***Figure S4.*** **Mediation detection frequency of the control analysis for the extinction of fear.** Exploratory mediation was carried out using a predictor and a mediator drawn from normally distributed random numbers to evaluate the rate of false positive detections in our dataset. Bar height represents the number of times a mediation effect for a specific ROI-pair surpassed the detection threshold when regressing each learning index onto the control predictor in a repeated cross-validation scheme with 1000 repeats. Results for functional- and structural connectivity mediators are indicated by grey and blue bars, respectively. ACC, dorsal anterior cingulate cortex; AMY, amygdala; CEB, cerebellar nuclei; HIP, hippocampus; PFC, ventromedial prefrontal cortex.

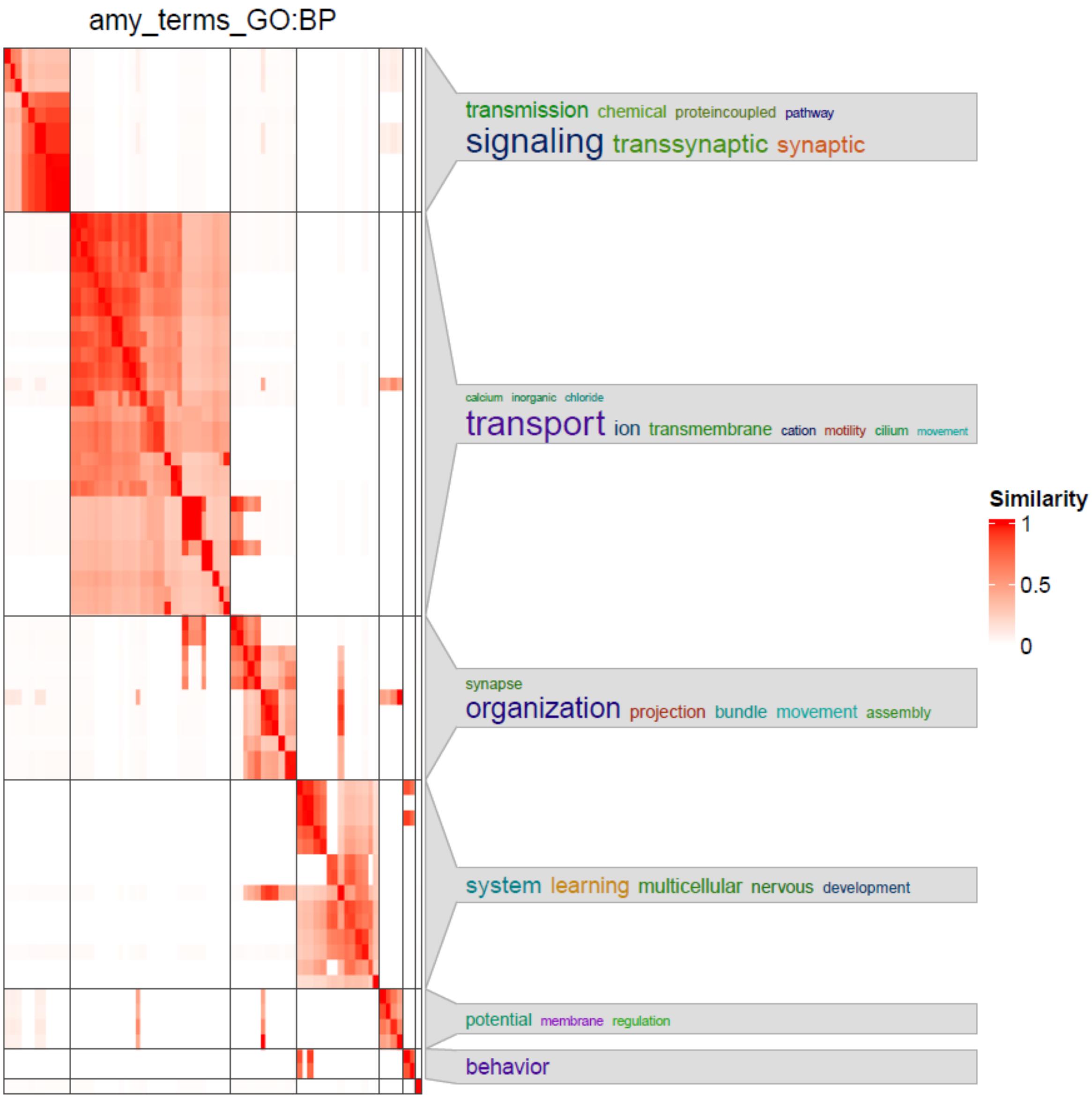

***Figure S5*. Similarity clusters of GO-terms that showed enrichment in the amygdala DCC-module.** Similar terms are clustered together. Intense colors indicate higher similarity. One keyword cloud with annotations summarizing enriched pathways is shown for each cluster. amy, amygdala; GO, gene ontology; BP, biological process.

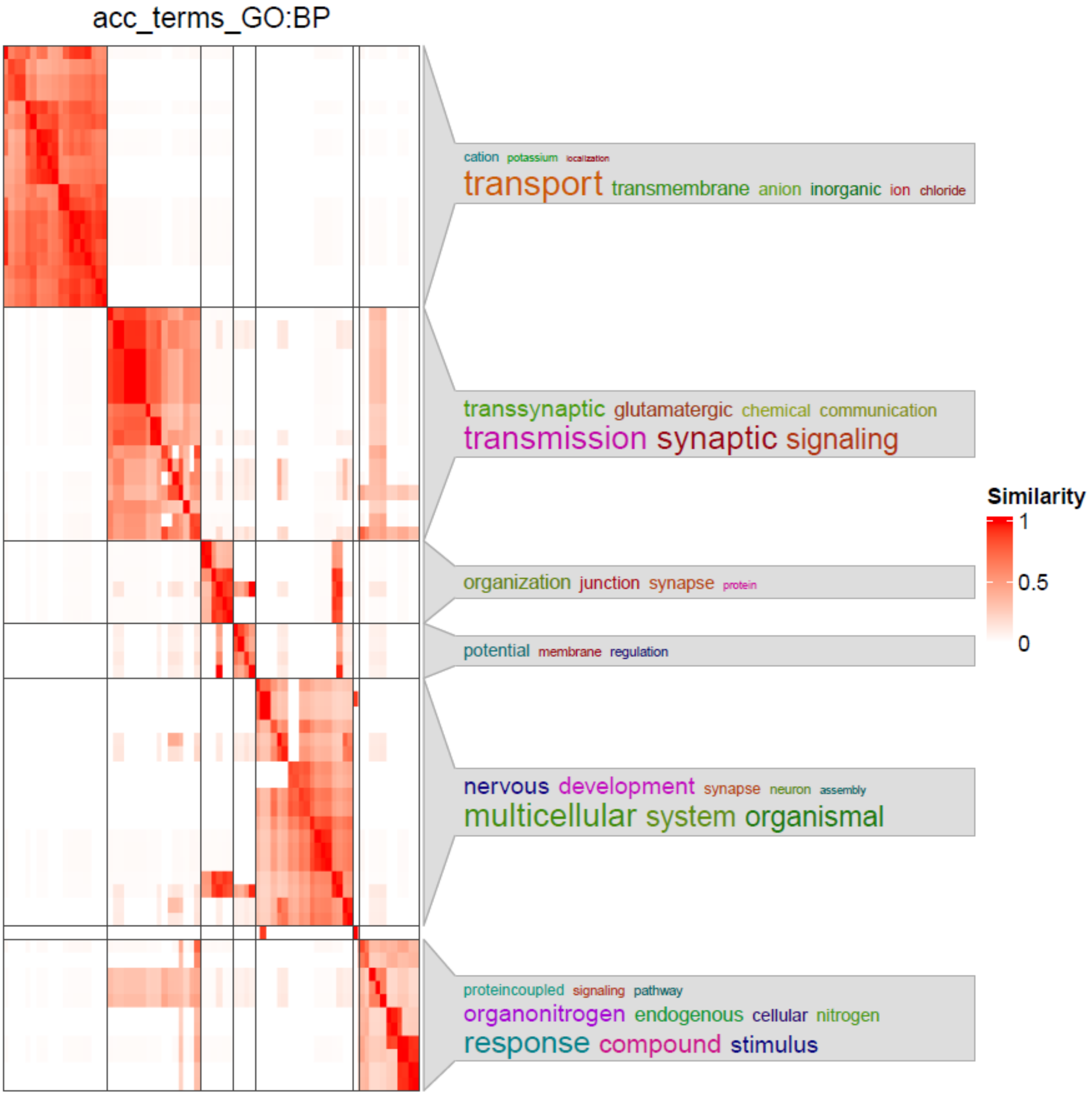

***Figure S6.*** **Similarity clusters of GO-terms that showed enrichment in the anterior cingulate DCC-module.** Similar terms are clustered together. Intense colors indicate higher similarity. One keyword cloud with annotations summarizing enriched pathways is shown for each cluster. acc, anterior cingulate; GO, gene ontology; BP, biological process.

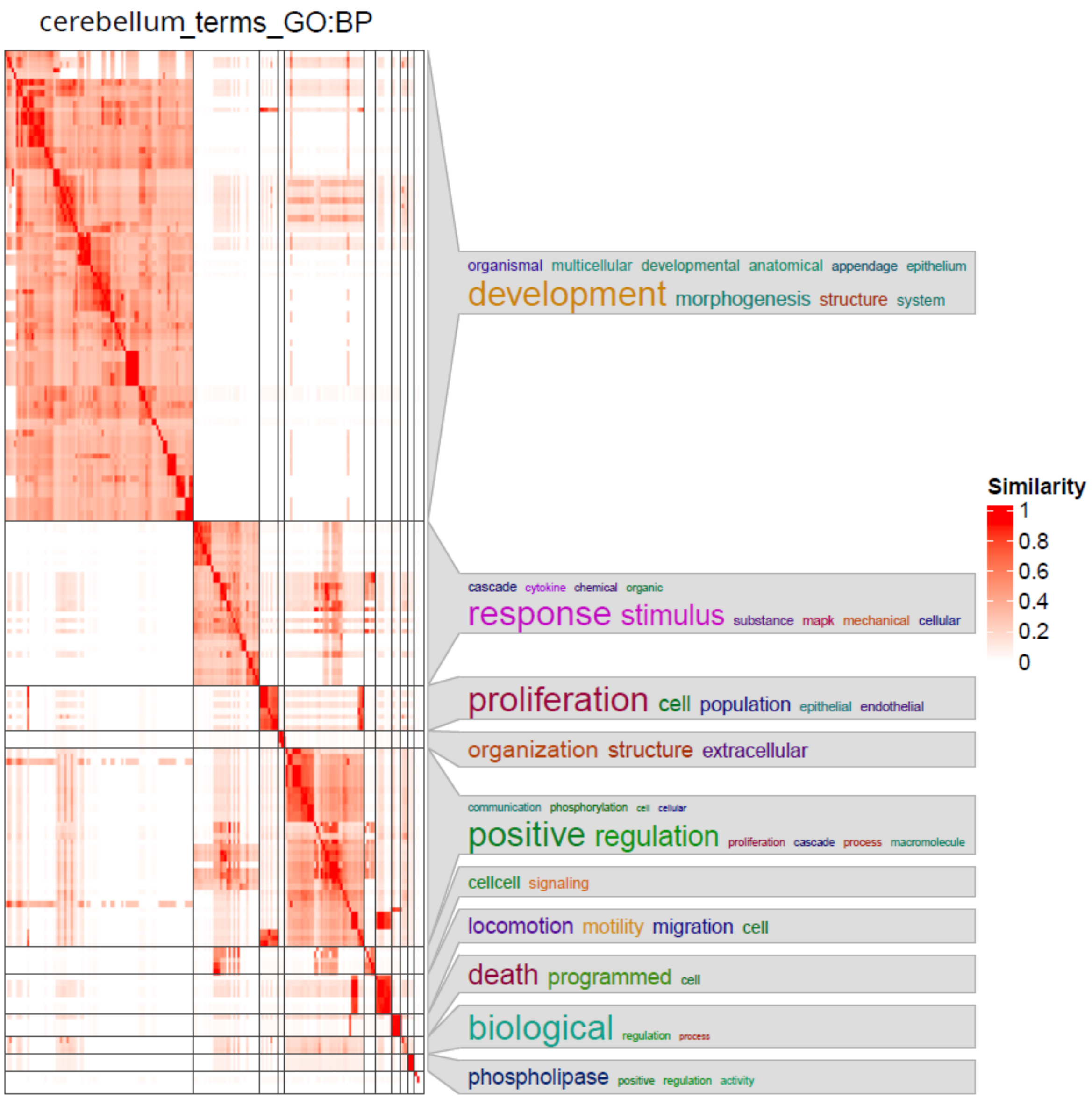

***Figure S7.*** **Similarity clusters of GO-terms that showed enrichment in the cerebellum DCC-module.** Similar terms are clustered together. Intense colors indicate higher similarity. One keyword cloud with annotations summarizing enriched pathways is shown for each cluster. GO, gene ontology; BP, biological process.

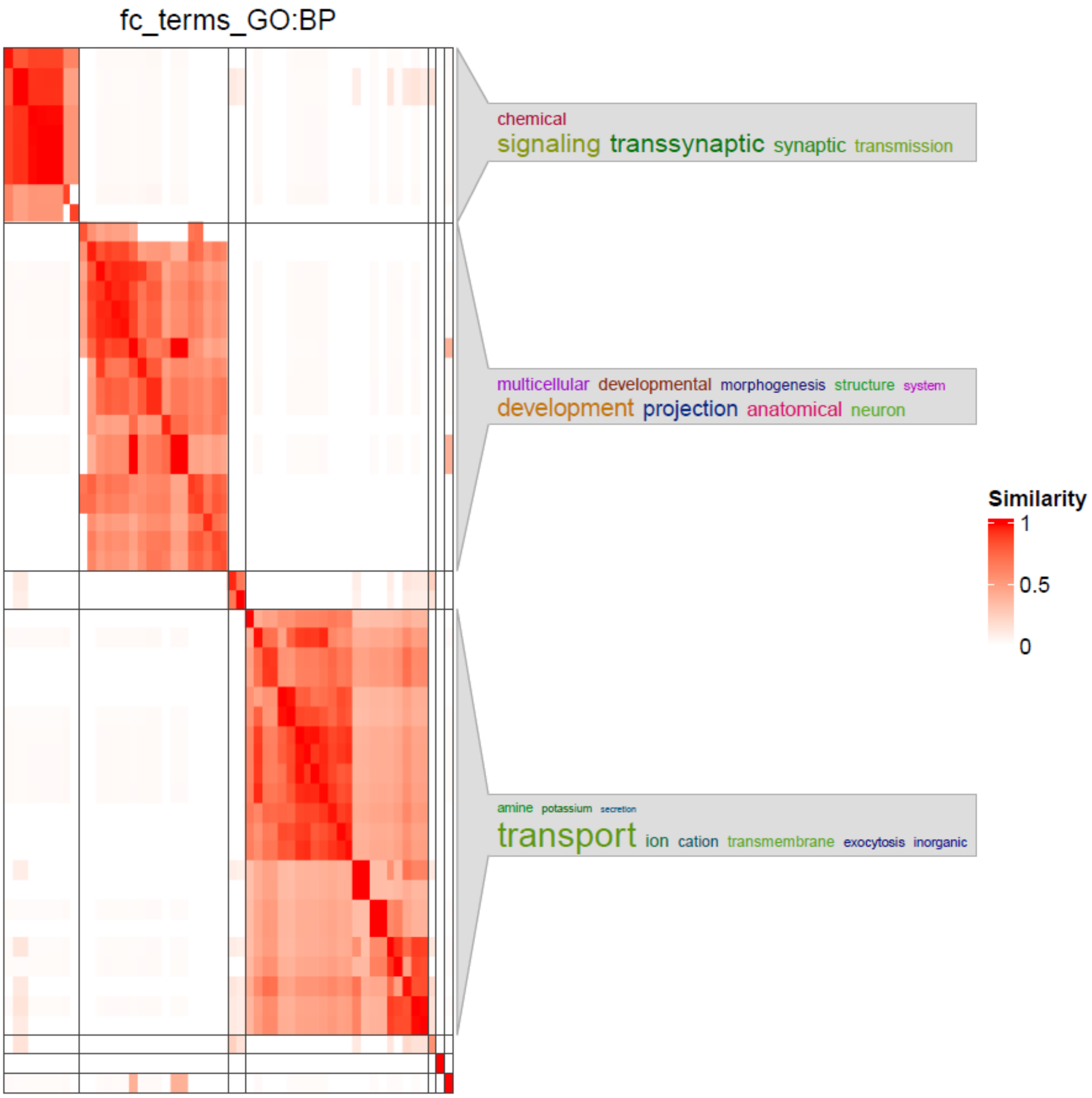

***Figure S8.*** **Similarity clusters of GO-terms that showed enrichment in the frontal cortex DCC-module.** Similar terms are clustered together. Intense colors indicate higher similarity. One keyword cloud with annotations summarizing enriched pathways is shown for each cluster. fc, frontal cortex; GO, gene ontology; BP, biological process.

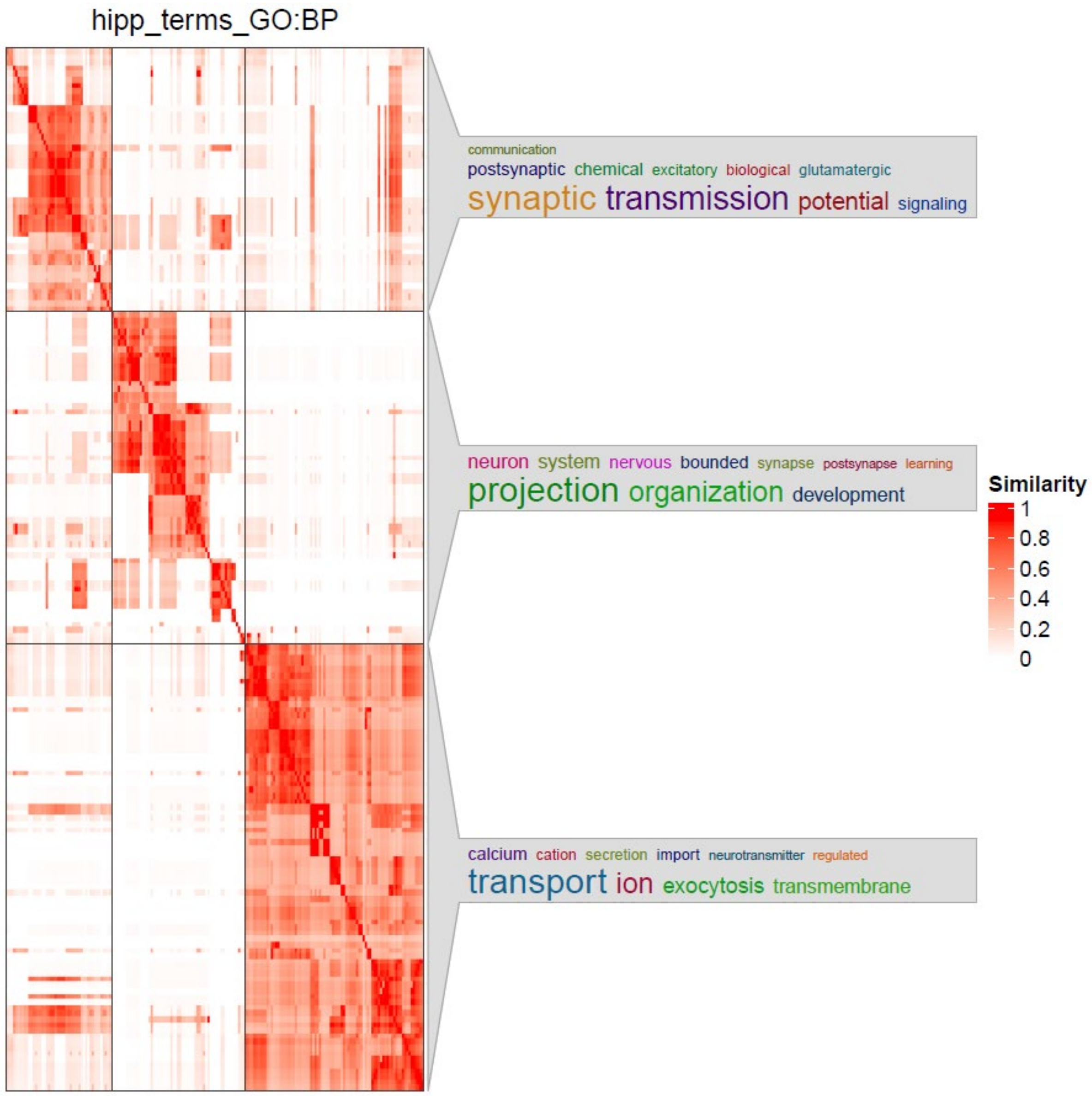

***Figure S9.*** **Similarity clusters of GO-terms that showed enrichment in the hippocampus DCC-module.** Similar terms are clustered together. Intense colors indicate higher similarity. One keyword cloud with annotations summarizing enriched pathways is shown for each cluster. hipp, hippocampus; GO, gene ontology; BP, biological process.

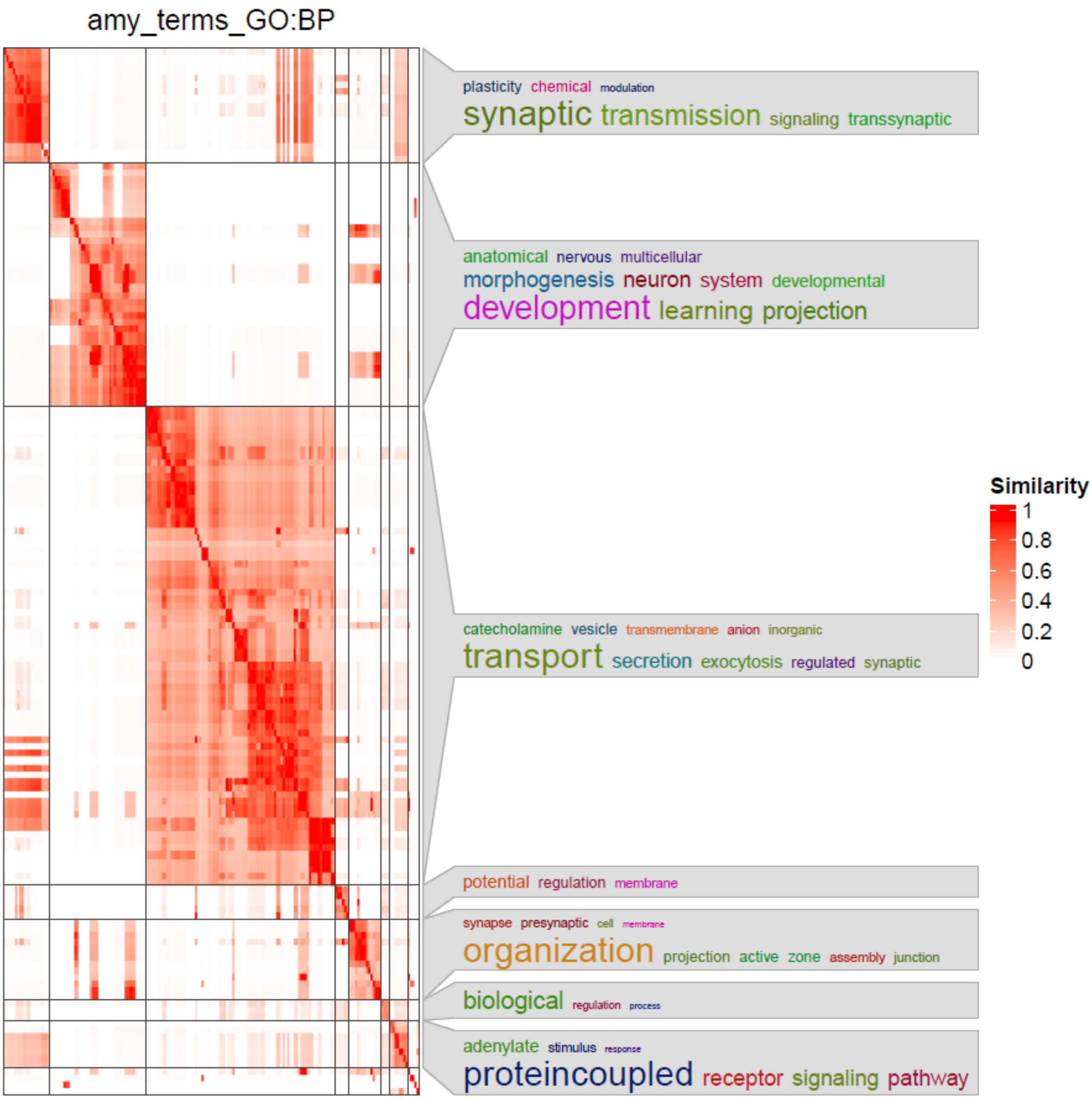

***Figure S10.*** **Similarity clusters of GO-terms that showed enrichment in the amygdala learning-module.** Similar terms are clustered together. Intense colors indicate higher similarity. One keyword cloud with annotations summarizing enriched pathways is shown for each cluster. amy, amygdala; GO, gene ontology; BP, biological process.

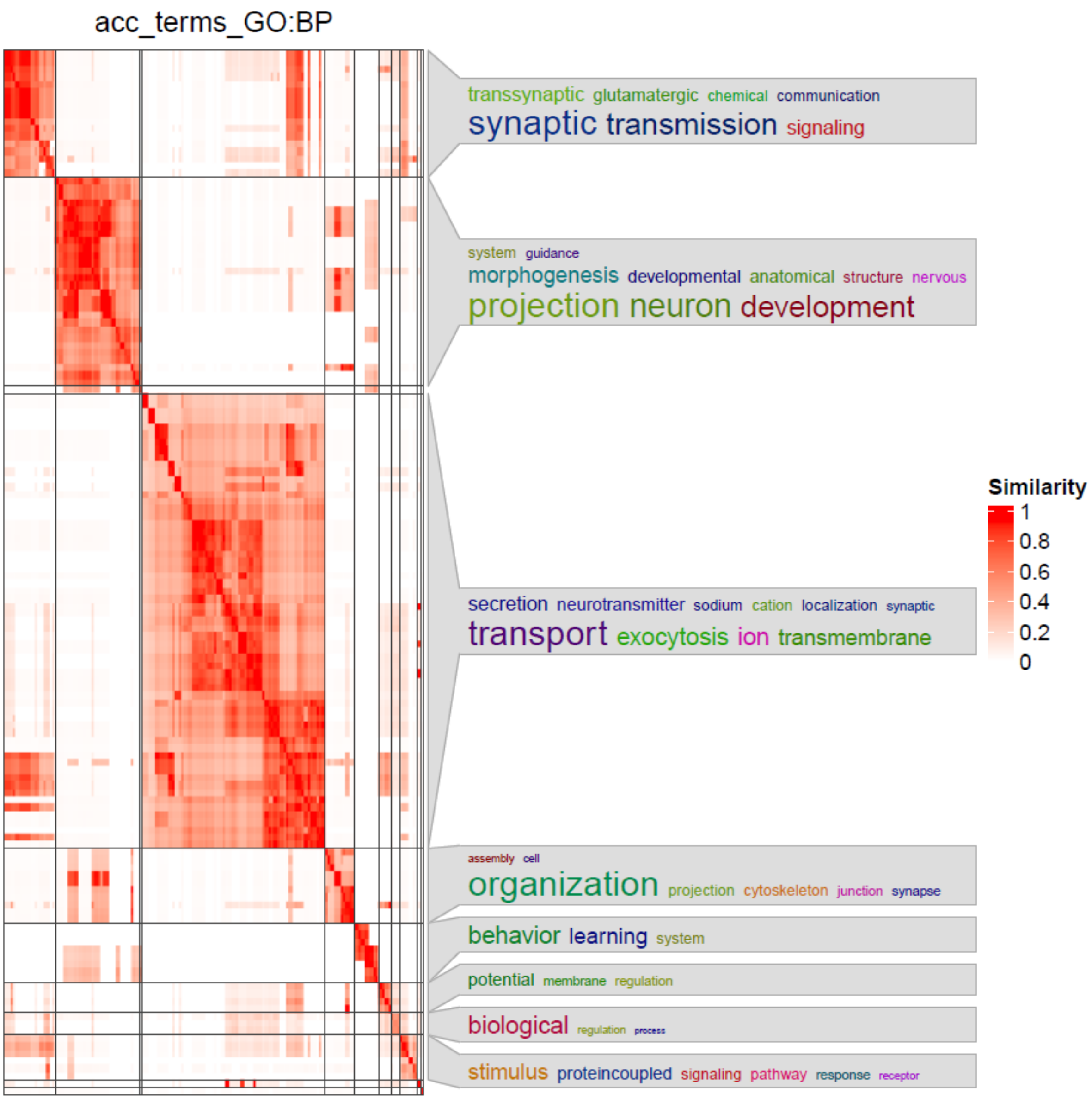

***Figure S11.*** **Similarity clusters of GO-terms that showed enrichment in the anterior cingulate learning-module.** Similar terms are clustered together. Intense colors indicate higher similarity. One keyword cloud with annotations summarizing enriched pathways is shown for each cluster. acc, anterior cingulate; GO, gene ontology; BP, biological process.

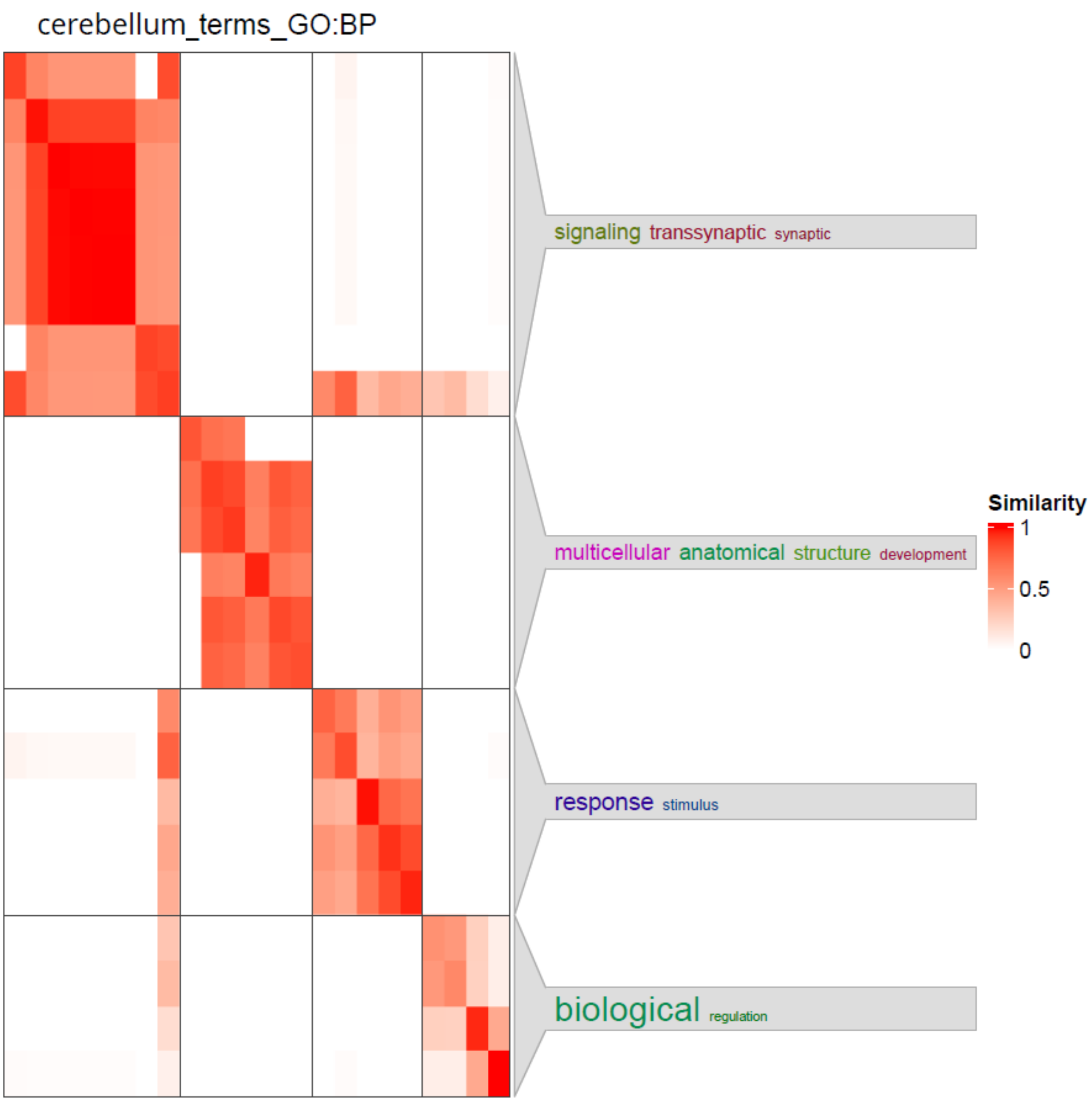

***Figure S12.*** **Similarity clusters of GO-terms that showed enrichment in the cerebellum learning-module.** Similar terms are clustered together. Intense colors indicate higher similarity. One keyword cloud with annotations summarizing enriched pathways is shown for each cluster. GO, gene ontology; BP, biological process.

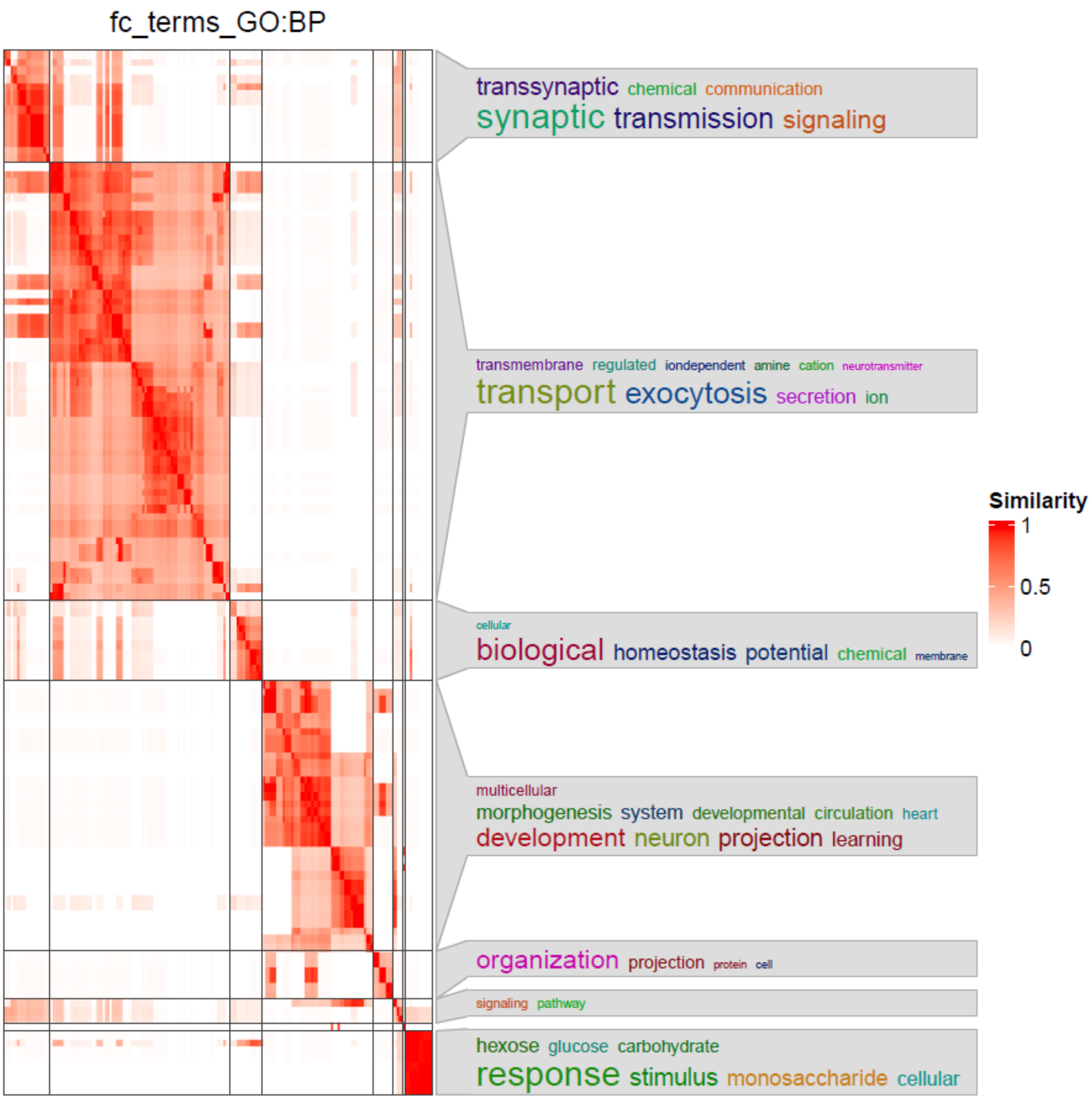

***Figure S13.*** **Similarity clusters of GO-terms that showed enrichment in the frontal cortex learning-module.** Similar terms are clustered together. Intense colors indicate higher similarity. One keyword cloud with annotations summarizing enriched pathways is shown for each cluster. fc, frontal cortex; GO, gene ontology; BP, biological process.

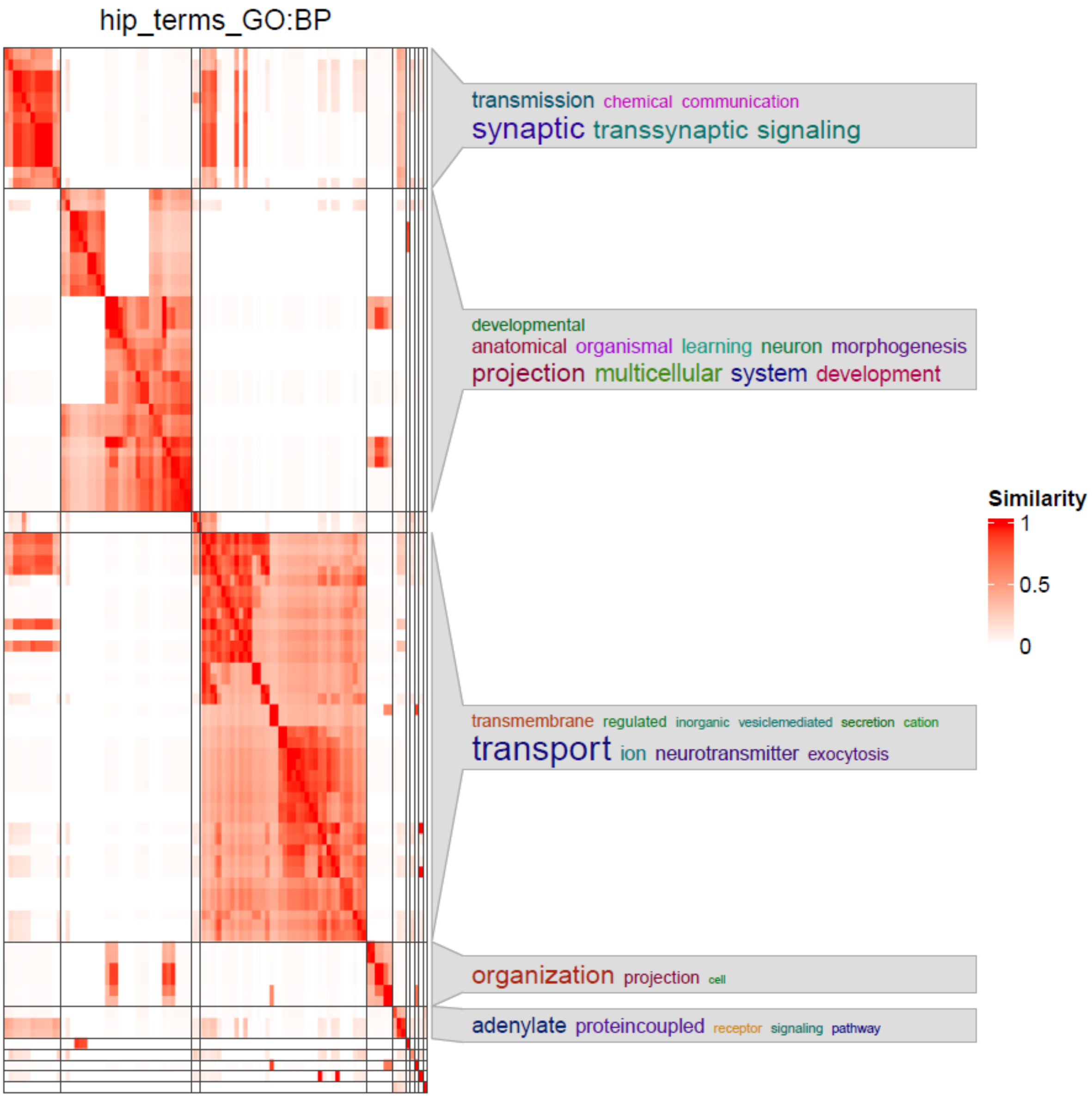

***Figure S14.*** **Similarity clusters of GO-terms that showed enrichment in the hippocampus learning-module.** Similar terms are clustered together. Intense colors indicate higher similarity. One keyword cloud with annotations summarizing enriched pathways is shown for each cluster. hipp, hippocampus; GO, gene ontology; BP, biological process.
